# The Mechanism of Vascular Endothelial Dysfunction Induced by Ferroptosis Mediated by NARFL Knockout

**DOI:** 10.1101/2024.02.06.24302421

**Authors:** Hui Hu, Jing Luo, Li Yu, Daoxi Qi, Boyu Li, Yating Cheng, Chen Wang, Xiaokang Zhang, Qiyong Lou, Gang Zhai, Yonglin Ruan, Jianfei Huang, Shengchi Shi, Zhan Yin, Fang Zheng

## Abstract

**BACKGROUND:** Nuclear prelamin A recognition factor-like (NARFL) plays a crucial role in cytosolic iron-sulfur protein assembly (CIA) and protects cells against oxidative stress. In our previous study, we identified a novel homozygous mutation in NARFL that led to decreased expression in a consanguineous family with diffuse pulmonary arteriovenous malformations (DPAVMs) secondary to pulmonary hypertension. Additionally, we observed that narfl deletion in zebrafish resulted in larvae lethality, subintestinal vessel malformation, and increased oxidative stress. In this study, we aimed to further investigate the function of NARFL and elucidate the pathological manifestations of NARFL deficiency in zebrafish models, cellular models, mouse models, and clinical samples, focusing on the underlying molecular mechanisms.

**METHODS:** We observed the behavioral and phenotypic abnormalities in zebrafish caused by narfl deletion and investigated the mechanism behind vascular morphological abnormalities. Furthermore, we constructed *NARFL* gene knockout stable cell lines in human pulmonary microvascular endothelial cells (HPMEC) to examine the morphological and functional changes in endothelial cells caused by NARFL deletion. We studied the effects of NARFL deletion on ferroptosis and its potential rescue using a ferroptosis inhibitor. To investigate the function of the human NARFL homolog Ciao3 gene in vascular development, we created a mouse model with a knockout of the *Ciao3* gene. Finally, we compared the distribution of tagSNPs of NARFL using the SNaPshot method between cases and controls to confirm the role of the Ciao3 gene in endothelial dysfunction.

**RESULTS:** Narfl deletion in zebrafish resulted in larvae lethality, vascular malformation with abnormal blood flow, abnormal blood-brain barrier (BBB) structure, and brain neuron lesions. Fluorescence probe detection showed increased iron, enhanced oxidative stress, lipid peroxidation, and decreased mitochondrial respiration in response to narfl deficiency, which could be partially alleviated by the use of the ferroptosis inhibitor Ferrostatin-1. We observed downregulation of the iron-sulfur protein cyp2p8 expression in blood vessels of narfl-deficient zebrafish through qRT-PCR and WISH experiments. In HPMEC cells, NARFL deficiency resulted in decreased proliferation, abnormal mitochondrial morphology, increased levels of iron and oxidative stress, and decreased mitochondrial respiration. Functional experiments on endothelial cells revealed decreased tube formation ability and enhanced permeability in response to NARFL deficiency. WB experiments showed downregulation of GPX4, SLC7A11, and Ferritin, while TFR1 and IRP1 were upregulated. Downregulation of NARFL also affected the expression of the iron-sulfur protein CYP2J2. Co-IP results indicated that NARFL deletion led to incompatibility among the CIA system-associated proteins. In mice, Ciao3 deletion in the embryonic stage resulted in embryonic death, vascular dysplasia, impaired differentiation of endothelial progenitor cells, and abnormalities in the expression of ferroptosis-related proteins. Reduction of Ciao3 impaired vascular function and decreased ring formation ability in adult heterozygous mice. *NARFL* polymorphisms rs11248948, rs2071952, and rs611289 were identified as susceptible sites for epilepsy, while rs11792680 was associated with susceptibility to pulmonary hypertension, epilepsy, and neurodegenerative diseases.

**CONCLUSION:** NARFL knockout disrupts its interaction with CIA system-related proteins, leading to decreased aconitase activity, increased IRP1 activity, endothelial cell ferroptosis pathway abnormalities, enhanced ferroptosis and oxidative stress, and ultimately vascular endothelial dysfunction. This dysfunction is responsible for the death of embryos in *narfl-/-* zebrafish and *Ciao3-/-* mice, as well as the susceptibility to pulmonary hypertension, epilepsy, and neurodegenerative diseases.

**What Is New?:**

1. Elucidation of the mechanism behind NARFL knockout-induced death through dynamic visualization experiments *in vivo* and mechanism and function experiments *in vitro:* The study explored the function of NARFL, as it is known as a “knockout lethal” protein. Both *in vivo* and *in vitro* experiments have confirmed that NARFL acts as the “transmitter” of cytoplasmic iron-sulfur clusters. Its absence prevents interaction with associated proteins of the CIA system, leading to reduced cisaconitase activity, enhanced IRP1 activity, ferroptosis of endothelial cells, and increased oxidative stress, eventually resulting in cell death.
2. Providing new research ideas for the study of cytoplasmic iron-sulfur proteins: Most current studies focus on the function of mitochondrial iron-sulfur proteins and their relationship with iron death. However, research on extramitochondrial iron-sulfur proteins is relatively limited. This study provides data support and research ideas for understanding the function of extramitochondrial iron-sulfur proteins by exploring the pathological mechanism of NARFL and the mediation of iron-sulfur protein maturation.

**What Are the Clinical Implications?:** From rare diseases to common diseases: Through the investigation of the lethal mechanism of NARFL knockout and the study of *NARFL* gene polymorphisms associated with vascular endothelial dysfunction diseases, we propose the hypothesis that NARFL may be a susceptibility gene for these diseases. This study provides data support for the hypothesis and contributes to our understanding of the role of NARFL in vascular endothelial dysfunction diseases.

## METHODS

The materials and methods that support the study findings are available from the corresponding author on reasonable request.

### Human and Animal Subjects and Ethical Considerations

In the case of patients with pulmonary hypertension involving pulmonary veins or capillaries, inclusion criteria required an average pulmonary arterial pressure (mPAP) of ≥ 25 mmHg measured by right cardiac catheterization or estimated from echocardiography using parameters such as pulmonary valve regurgitation beam spectrum, right atrial regurgitation beam spectrum, and tricuspid regurgitation flow. Exclusion criteria included pulmonary hypertension caused by congenital heart disease, left ventricular disease, lung disease, and/or hypoxia. For patients with cerebral small vascular disease, inclusion criteria encompassed various cerebrovascular diseases such as epilepsy, Alzheimer’s disease, lacunar cerebral infarction, Binswanger encephalopathy, autosomal dominant cerebral arteriopathy, and amyloidosis cerebrovascular disease with subcortical cerebral infarction and leukoencephalopathy. MRI findings needed to meet the imaging diagnostic criteria of cerebrovascular disease, including lacunar cerebral infarction, white matter changes, and cerebral microhemorrhage. Exclusion criteria encompassed imaging changes caused by carbon monoxide poisoning, severe sleep apnea syndrome, infection-related brain changes, and trauma-induced brain changes. For patients with systemic lupus erythematosus due to endothelial cell injury, clinical symptoms needed to meet the diagnostic criteria revised by the American Rheumatology Society in 1997. Immunological abnormalities included positivity for anti-ds-DNA antibodies, anti-Sm antibodies, or anti-phospholipid antibodies (which encompassed indicators such as anticardiolipin antibody, positive lupus anticoagulant, or false-positive syphilis serum test results persisting for at least 6 months). Additionally, elevated levels of markers related to systemic lupus erythematosus endothelial cell injury, such as vWF, MDA, GSH, and SOD, were observed.Patient information, including age, sex, smoking and alcohol consumption status, body mass index (BMI), echocardiography results, and routine biochemical indicators, was collected by consulting enrollment and admission records, electronic medical records, and laboratory examination information. All data were collected using a blind method and collected, organized, entered, and verified by different personnel. This study strictly adhered to the principles outlined in the Helsinki Declaration and received approval from the Ethics Committee of Zhongnan Hospital, Wuhan University.

### Statistical Analysis

Mean±SEM or mean±SD was used to represent the data. For cell culture data, three independent experiments were performed in triplicate. Animal numbers were determined based on the calculation of a ≥20% difference between the means of experimental and control groups with a statistical power of 80% and a standard deviation (SD) of 10%. The normality of data was confirmed using Shapiro-Wilk testing. For comparisons between two groups with normally distributed data, a two-tailed Student’s t-test was performed. When comparing multiple groups, either one-way or two-way ANOVA was used, as appropriate. A p-value of less than 0.05 was considered statistically significant.

### Background

In a consanguineous family with diffuse pulmonary arteriovenous malformations (DPAVMs) leading to pulmonary arterial hypertension, the research group identified a novel homozygous mutation (pSer161Ile) in the NARFL gene^1^(**Figure 1A-E in the online-only Data Supplement**). This mutation was associated with a decrease in mRNA stability and expression levels of NARFL, and narfl-/- zebrafish embryos exhibited lethal vascular malformation, indicating a potential involvement of NARFL in the development of pulmonary arteriovenous malformation.

**Figure 1:**
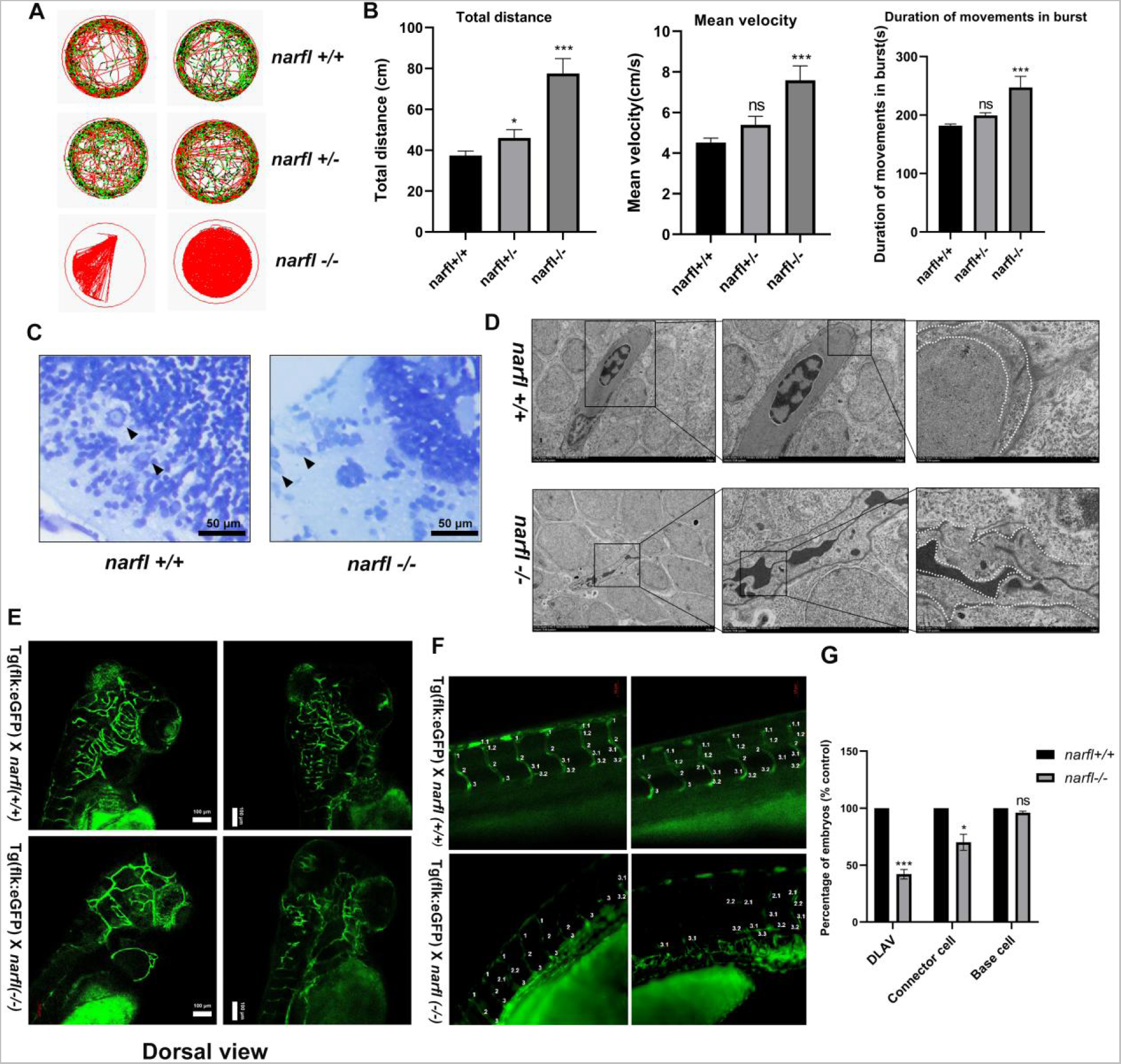
Narfl Deficiency Results in Abnormal Behavior, Abnormal Blood Vessels, and Neurons in Zebrafish. (**A**) Swimming behavior trajectories of zebrafish with different genotypes under normal lighting conditions. (**B**) Swimming behavior analysis of zebrafish with different genotypes within 60 minutes under normal lighting, including total swimming distance, average swimming speed, and burst duration. (**C**) Toluidine blue staining was performed to assess Nissl body morphology in the brains of zebrafish with different genotypes. (**D**) Transmission electron microscopy (TEM) was used to observe the ultrastructure of the blood-brain barrier in the brains of zebrafish with different genotypes at magnifications of 1500×, 5000×, and 10000×, respectively. (**E**) Fluorescence inverted microscope imaging system (dorsal view) was utilized to observe the cerebral vascular morphology of zebrafish with different genotypes. (**F**) Fluorescence confocal microscopy imaging and quantitative analysis of zebrafish vascular segments revealed the absence of junction cells in the *narfl-/-* dorsal longitudinal anastomosis. (**G**) Disordered or absent connective cells were observed in the *narfl-/-* zebrafish dorsal longitudinal anastomosis vessels, along with disordered and distorted structures of the dorsal aorta and PCV.

The CIA system, which operates in the eukaryotic cytoplasm, comprises various proteins that work together to perform ISC-related functions^2,3,7,8^. Iron-sulfur proteins containing iron-sulfur clusters (ISC) serve as prosthetic groups for electron transport proteins and active groups for enzymes, participating in essential physiological processes such as energy metabolism, amino acid and iron metabolism, DNA replication and repair, and gene expression regulation^4^. While the process of ISC core biogenesis has been extensively studied, particularly in mitochondrial iron-sulfur proteins, research on the function of iron-sulfur proteins outside the mitochondria remains limited^4–6^.

The CIA system, which operates in the eukaryotic cytoplasm, comprises various proteins that work together to perform ISC-related functions. These include nucleotide binding protein 1 (NUBP1) and nucleotide binding protein 2 (NUBP2) as scaffold proteins that bind and accept ISC, NARFL as an intermediate carrier protein for transmitting ISC, and a CIA targeting complex (CTC) consisting of CIA Component 1 (CIAO1), Methyl methanesulfonate sensitivity 19 (MMS19), and MIP18 (MMS19-interacting protein of 18kDa or CIAO2/FAM96B), which inserts ISC into specific apoproteins^9, 10^. NARFL, as an essential component of CIA, influences the synthesis and maturation of cytoplasmic iron-sulfur proteins. Notably, cytoplasmic aconitase (ACO1), a well-studied cytoplasmic iron-sulfur protein, plays a critical role in catalyzing isocitric acid. ACO1 functions as ACO1 when it receives ISC^11,12^. Upon ISC loss, it transforms into iron regulatory protein 1 (IRP1), acting as an apoprotein of ACO1^13,14^. Consequently, the knockdown of NARFL may lead to decreased ACO1 activity, increased IRP1 protein activity, and disruptions in iron metabolism, potentially contributing to vascular dysfunction.

## RESULTS

### Narfl Deficiency Leads to Abnormal Behavior and Abnormal Blood Vessels and Neurons in Zebrafish

The swimming trajectories of different zebrafish genotypes were analyzed under normal illumination for a duration of 60 minutes (**Figure 1A**). The results revealed significant differences in total swimming distance, average swimming speed, and outbreak duration between *narfl-/-* zebrafish and wild-type zebrafish (**Figure 1B**). Notably, *narfl-/-* zebrafish exhibited increased activity, often displaying spontaneous and irregular movements reminiscent of epileptic seizures^15,16^. Seeking to elucidate the underlying cause of this phenotype, toluidine blue staining of 7-day-old zebrafish revealed dissolved Nissl corpuscles, flattened morphology, and shifted nuclei in *narfl-/-* zebrafish compared to wild-type zebrafish (**Figure 1C**), indicating pathological changes in neurons. Examination of the blood-brain barrier (BBB) ultrastructure using transmission electron microscopy showed that endothelial cells of *narfl-/-* zebrafish had evident shrinkage, expansion, and basement membrane breaks, in contrast to the plump and intact structure observed in wild-type zebrafish (**Figure 1D**). TUNEL staining of the brains of 9-day-old zebrafish did not reveal significant differences (**Figure 2C in the online-only Data Supplement**). Furthermore, by establishing hybridizations between *Tg (flk: eGFP)* zebrafish models and *narfl (+/+)* or *narfl (-/-)*, confocal microscopy observations of cerebral vessels demonstrated defects in *narfl (-/-)* zebrafish, characterized by decreased vessel quantity and disorganized arrangement (**Figure 1E and Figure 2B in the online-only Data Supplement**). Previous studies have shown that narfl deletion resulted in early zebrafish embryo death17. Notably, the growth of *narfl -/-* zebrafish embryos displayed various deformities and irregularities (**Figure 2A in the online-only Data Supplement**). Further analysis using fluorescence confocal microscopy and quantitative assessment of zebrafish vascular segments revealed disordered or absent connective cells in *narfl -/-* zebrafish dorsal longitudinal anastomosis vessels, along with notable structural disorganization and distortion of the dorsal aorta and posterior cardinal vein (PCV) (**Figure 1F and G and Figure 2D in the online-only Data Supplement**). Detection of γ-H2AX indicated substantial DNA damage in the dorsal aorta and PCV (**Figure 2E and F in the online-only Data Supplement**).

**Figure 2:**
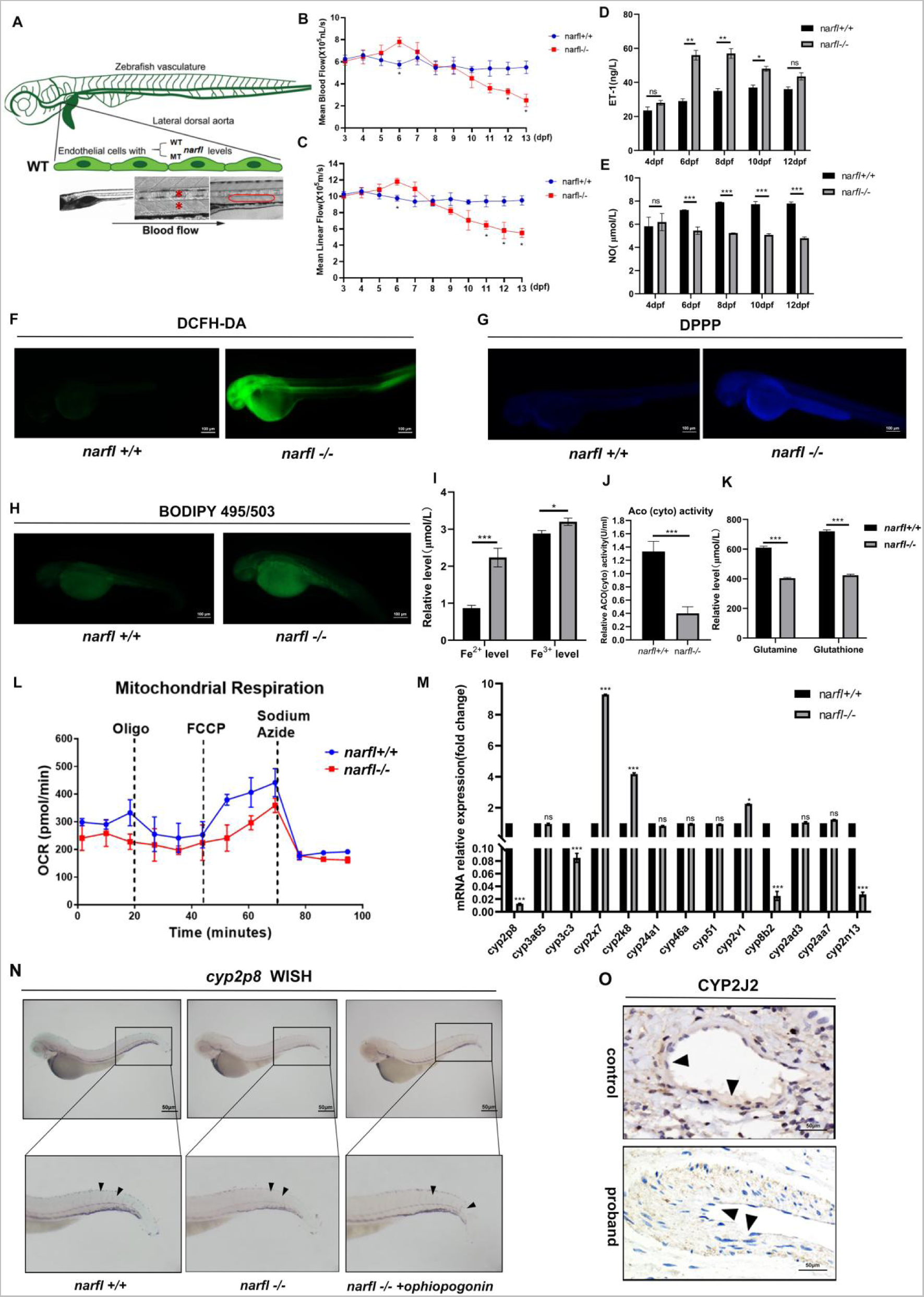
Narfl Deficiency Activates Zebrafish Endothelial Dysfunction by Upregulating Iron Level, Reactive Oxygen Species Production, and Lipid Peroxidation. (**A**) The Micro Zebra Lab system detected the blood flow of zebrafish, with the red asterisk denoting line speed and the red box denoting average speed. (**B-E**) Mean blood flow velocity (**B, D**) and mean linear velocity (**C, E**) of *narfl-/-* zebrafish were compared to wild-type zebrafish at different developmental stages. Significant differences were observed at specific time points. (**F**) The DCFH-DA probe was utilized to measure oxidative stress in 5 dpf zebrafish. Stronger green fluorescence indicated higher oxidative stress levels. Quantitative analysis showed enhanced fluorescence intensity in *narfl-/-* zebrafish compared to wild type. (**G**) The DPPP probe was used to detect lipid peroxidation levels in 5 dpf zebrafish. Stronger purple fluorescence indicated higher lipid peroxidation levels. (**H**) The BODIPY 493/503 probe detected neutral lipid levels in zebrafish, with stronger green fluorescence indicating higher lipid levels. (**I**) Iron levels in zebrafish were measured by a colorimetric method, showing significantly higher iron levels in *narfl-/-* zebrafish compared to wild type. Fe^3+^ levels were also increased. (**J**) Cytoplasmic cisaconitase activity was assessed in zebrafish, with significantly enhanced activity observed in *narfl-/-* zebrafish. (**K**) Glutathione and glutamine levels in zebrafish were measured, revealing significantly reduced levels in *narfl-/-* zebrafish. (**L**) Mitochondrial respiration integration of wild-type and *narfl-/-* zebrafish demonstrated a decrease in mitochondrial respiratory function in *narfl-/-* zebrafish. (**M**) qRT-PCR was used to verify related genes of P450 family with differences in RNA sequencing, and the results showed that *cyp2p8, cyp3c3, cyp8b2* and *cyp2n13* were significantly decreased in *narfl-/-* zebrafish. *cyp2x7, cyp2k8* and *cyp2v1* were significantly increased, while there was no significant difference in other genes. (**N**) In situ hybridization detected cyp2p8 expression in 5 dpf wild-type and *narfl-/-* zebrafish, as well as after treatment with the cyp2p8-specific activator ophiopogon D. The locations indicated by the black arrows showed the sites with positive *cyp2p8* probe signals (**O**) Immunohistochemistry was performed to analyze the expression of CYP2J2 in adjacent lung tissue and in progenitors with diffuse pulmonary malformation. CYP2J2 expression in the cytoplasm was reduced in progenitors with down-regulated NARFL expression.

### Narfl Deficiency Induces Zebrafish Dysangiogenesis by Upregulating Iron Levels and ROS Production and Lipid Peroxidation

To investigate whether the abnormal vascular morphology in *narfl-/-* zebrafish is a result of endothelial cell dysfunction and hemodynamic abnormalities, we utilized the Micro Zebra Lab system from the 3 dpf of zebrafish development (**Figure 2A**). Considering the high mortality rate prior to the 13 dpf, we monitored the zebrafish until the end of the 13 dpf day. Subsequently, we calculated the mean blood flow velocity and mean linear velocity from 3 dpf to 13 dpf. The statistical analysis revealed that the mean blood flow velocity and linear velocity in *narfl-/-* zebrafish showed no significant differences compared to wild-type zebrafish at 3-5 dpf, but were significantly higher at 6 dpf, significantly lower at 7-11 dpf, and significantly lower at 11-13 dpf **(Figure 2B, C**).

To assess the levels of endothelin-1 (ET-1) and nitric oxide (NO), we measured their concentrations at 4 dpf, 6 dpf, 8 dpf, 10 dpf, and 12 dpf. The results revealed that the levels of ET-1 in *narfl-/-* zebrafish at 6 dpf, 8 dpf, and 10 dpf were significantly higher than those in wild-type zebrafish. Conversely, the levels of NO at 6 dpf, 8 dpf, 10 dpf, and 12 dpf were significantly lower in narfl-/- zebrafish compared to wild-type zebrafish (**Figure 2D, E**).

To assess reactive oxygen species (ROS) levels, we utilized the DCFH-DA probe in zebrafish (**Figure 2F**). The fluorescence intensity in *narfl-/-* zebrafish was significantly higher compared to wild-type zebrafish, indicating an elevated oxidative stress due to narfl deletion. We also employed the DPPP probe (**Figure 2G**) and the BODIPY 493/503 probe (**Figure 2H**) to measure lipid peroxidation in zebrafish. The results demonstrated that the fluorescence intensity was significantly higher in *narfl-/-* zebrafish compared to wild-type zebrafish, indicating an increase in lipid peroxidation due to narfl deletion.

However, the fluorescence intensity of apoptosis, as detected by AO staining, did not significantly increase in *narfl-/-* zebrafish compared to wild-type zebrafish (**Figure 3A in the online-only Data Supplement**), suggesting that narfl deletion did not significantly affect apoptosis. Using a colorimetric method, we measured the Fe^2+^ and Fe^3+^ contents in *narfl-/-* zebrafish and found that they were significantly higher compared to wild-type zebrafish (**Figure 2I**). Prussian blue staining of the zebrafish brain revealed a significant increase in hemosiderin content in *narfl-/-* zebrafish compared to wild-type zebrafish (**Figure 3B in the online-only Data Supplement**), indicating an elevation of iron levels in narfl deficiency.

**Figure 3:**
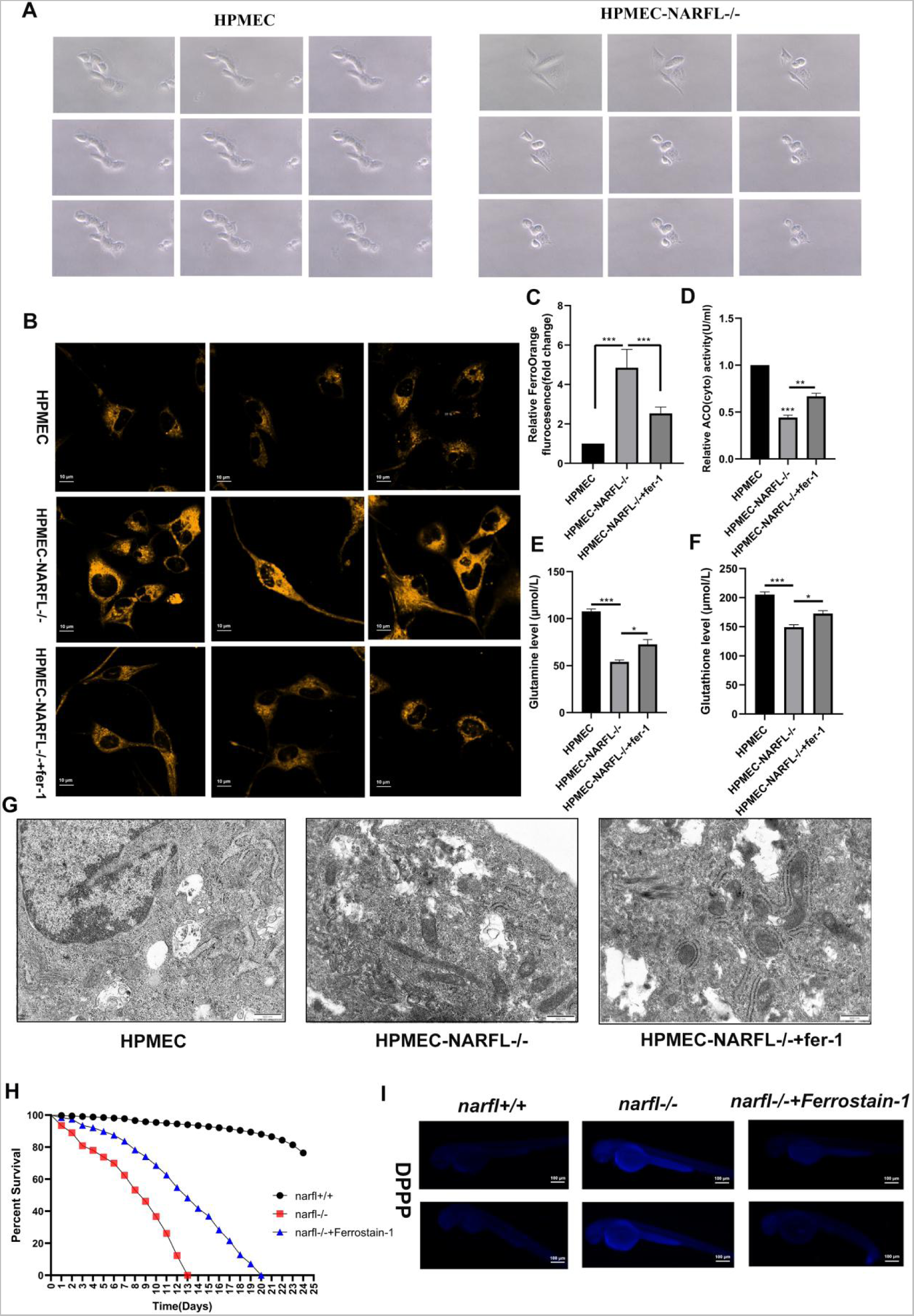
Endothelial Knockdown of NARFL Promotes Ferroptosis and Alleviation of Oxidative Stress Injury by Ferrostain-1. (**A**) Dynamic observation under a microscope and daily photography of HPMEC wild-type cells and *NARFL* mutant cells revealed significant differences in cell death patterns. *NARFL* mutant cells exhibited noticeable morphological changes from long and narrow to round, resembling cells undergoing ferroptosis. (**B**) Confocal fluorescence microscopy with FerroOrange probe showed fluorescence intensity in HPMEC, *NARFL-/-* HPMEC, and *NARFL-/-* HPMEC cells treated with Ferrostain-1 (6μM). Darker orange color indicated higher ferrous ion content in the cells. (**C**) Quantitative results of ferrous ion content measured by FerroOrange fluorescence probe showed a significant increase in HPMEC after NARFL deletion, which decreased after Ferrostain-1 addition. (**D-F**) Quantitative results of cytoplasmic cisaconitase (**D**), glutamine (**E**), and glutathione (**F**) in HPMEC, *NARFL-/-* HPMEC, and 6μM Ferrostain-1 treated *NARFL-/-* HPMEC cells illustrated decreased cytoplasmic cis-aconitase activity, intracellular glutathione, and glutamine content in endothelial cells after NARFL deletion. Partial restoration was observed upon treatment with the ferroptosis inhibitor Ferrostain-1. (**G**) Transmission electron microscopy of *NARFL-/-* HPMEC revealed distinct morphological differences in mitochondria compared to wild-type cells, exhibiting smaller size, increased membrane density, and reduced cristae, consistent with ferroptosis morphology.

In line with these findings, cytoplasmic cis-aconitase activity significantly increased in *narfl-/-* zebrafish (**Figure 2J**), while glutathione and glutamine (GSH-GL) content significantly decreased (**Figure 2K**).

### Narfl Deficiency Impairs Mitochondrial Respiratory Function and Downregulates Cyp2p8 Expression in Zebrafish

To assess the impact of narfl deletion on mitochondrial respiratory function in zebrafish, we used the Seahorse XFe24 cell metabolic respiratory dynamic analyzer to measure the changes in oxygen consumption rate (OCR) upon treatment with oligomycin, FCCP (a mitochondrial oxidative phosphorylation uncoupler), rotenone, and sodium azide. OCR reflects mitochondrial electron transfer and provides insights into respiratory function. We examined mitochondrial respiratory function in 11 wild-type and 8 *narfl-/-* zebrafish. Integration of the results revealed a significant impairment in mitochondrial respiratory function in *narfl-/-* zebrafish compared to wild-type zebrafish, indicating that narfl deletion leads to reduced mitochondrial respiratory function (**Figure 2L**).

In transcriptome sequencing analysis of *narfl +/+* and *narfl -/-* zebrafish, we found significant differences in a large number of genes related to iron metabolism. Among them, cytochrome P450 (CYP450) family genes, including *cyp2p8, cyp3a65, cyp3c3, cyp2x7, cyp2k8, cyp24a1, cyp46a, cyp51, cyp2v1, cyp8b2, cyp2ad3, cyp2aa7,* and *cyp2n13* exhibited significant downregulation in narfl-/- zebrafish (**Figure 3E in the online-only Data Supplement**). Notably, *cyp2p8* displayed the most significant downregulation and was chosen as the target downstream gene (**Figure 2M**). In situ hybridization with a *cyp2p8* probe on 5-dpf embryos revealed expression of *cyp2p8* in zebrafish blood vessels, with slightly enhanced signal in *narfl-/-* zebrafish treated with Ophiopogonin D, a specific activator of cyp2p8 (**Figure 2N**).

In humans, *cyp2p8* is known as *CYP2J2*. To examine whether NARFL deletion also leads to decreased expression of corresponding iron-sulfur proteins in humans, we performed immunohistochemical staining on lung tissues from the proband with NARFL downregulation and control samples (**Figure 2O**). The results showed that CYP2J2 expression, mainly in the cytoplasm, was decreased in the proband with downregulated NARFL expression, consistent with the findings in zebrafish. Thus, these results suggest that narfl deletion leads to downregulation of cytoplasmic iron-sulfur protein CYP2J2 expression in zebrafish, providing insights into the molecular mechanisms underlying Narfl deficiency-induced mitochondrial dysfunction and dysregulation of iron-sulfur metabolism.

### Endothelial Cell Knockdown of NARFL Promotes Ferroptosis and Ferrostain-1 can Alleviate Oxidative Stress Injury Caused by NARFL deficiencies

Immunofluorescence staining of lung tissues from a patient with pulmonary hypertension secondary to diffuse pulmonary arteriovenous malformation revealed a significant decrease in NARFL expression in the blood vessels compared to normal lung tissues. In addition, the expression of CD31, an endothelial cell marker, was decreased, while the expression of α-smooth muscle actin (α-SMA) was increased (**Figure 1F in the online-only Data Supplement**). To further investigate the mechanism, a NARFL knockout model was established using Human Pulmonary Microvascular Endothelial Cells (HPMECs) (**Figure 4A-D in the online-only Data Supplement**). The *NARFL-/-* HPMECs exhibited slow growth, increased cell death, and distinct morphological changes compared to wild-type cells. The morphology of NARFL mutant cells resembled that of cells undergoing ferroptosis. Furthermore, the proliferation ability of NARFL knockout cells was significantly decreased compared to wild-type cells (**Figure 4E in the online-only Data Supplement**).

**Figure 4:**
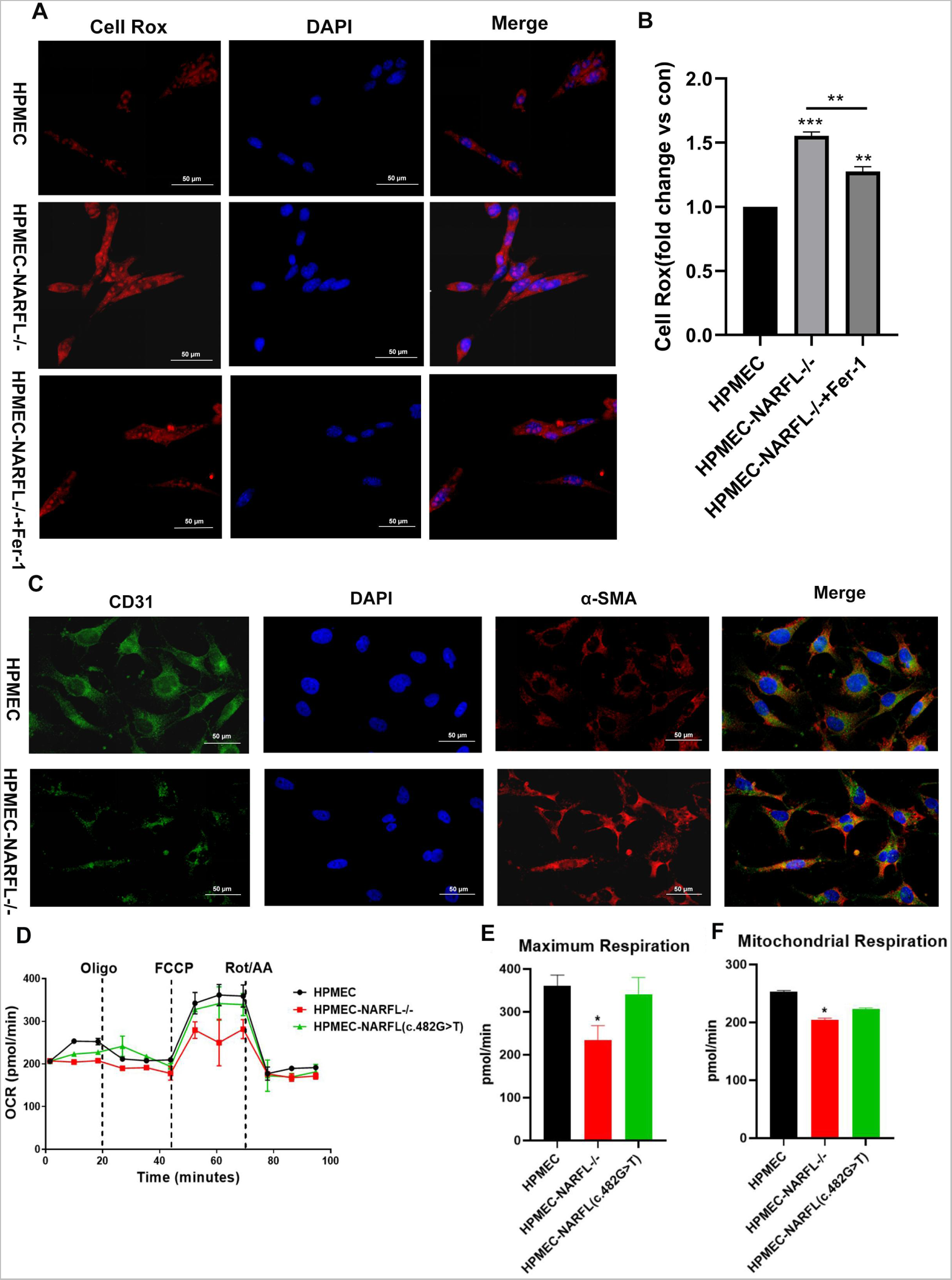
NARFL Gene Deletion Leads to Endothelial Cell Dysfunction. (**A**) Cell Rox kit was utilized to assess the oxidative stress level of endothelial cells. The intensity of red fluorescence indicates the level of cellular oxidative stress. (**B**) Quantitative analysis of cellular oxidative stress using CellRox kit revealed that NARFL deletion increased the level of oxidative stress in endothelial cells. Treatment with the ferroptosis inhibitor Ferrostain-1 partially alleviated the oxidative stress injury caused by NARFL gene deletion. (**C**) Immunofluorescence was performed to examine the expression levels of the endothelial cell marker CD31 and the fibroblast marker α-SMA in HPMEC and *NARFL-/-* HPMEC. Green fluorescence indicates CD31 expression, while red fluorescence indicates α-SMA expression. The results demonstrated that NARFL gene deletion downregulated CD31 and upregulated α-SMA expression. (**D-F**) Mitochondrial respiratory function curves of HPMEC, *NARFL-/-* HPMEC, and HPMEC transfected with *NARFL (c.482 G>T)* were generated. The black curve represents the wild-type cells, the green curve represents NARFL-transfected cells with the c.482 G>T point mutation, and the red curve represents HPMEC with NARFL deletion mutations. The results showed that mitochondrial respiratory function decreased in the presence of the *NARFL (c.482 G>T)* point mutant, although the difference compared to wild-type cells was minimal. On the other hand, NARFL deletion resulted in a significant decrease in mitochondrial function. (**H**) Survival analysis showed that 8μM Ferrostain-1 extended the survival time of *narfl-/-* zebrafish from 13 days to 21 days. (**I**) DPPP fluorescent probe-based detection of 5 dpf zebrafish treated with 8 μM Ferrostain-1 demonstrated inhibition of lipid peroxidation levels caused by narfl deletion.

Transmission electron microscopy analysis revealed altered mitochondrial morphology in NARFL mutant cells, characterized by smaller mitochondria, increased mitochondrial membrane density, and reduced cristae, resembling the morphology observed in ferroptosis (**Figure 3G and Figure 4F in the online-only Data Supplement**). To explore whether ferroptosis inhibitors can alleviate oxidative stress injury induced by NARFL deletion, Ferrostain-1 (a ferroptosis inhibitor) and α-Vitamin E (an oxidative stress inhibitor) were employed (**Figure 3C-D in the online-only Data Supplement**). The results demonstrated that 8 μM of Ferrostain-1 had the most pronounced effect on *narfl-/-* zebrafish, extending their survival time from 13 to 21 days (**Figure 3H**) and reducing lipid peroxidation levels caused by narfl deletion **(Figure 3I**). FerroOrange fluorescence results showed a significant increase in cellular ferrous ion content after NARFL deletion, which was significantly reduced upon addition of Ferrostain-1 (**Figure 3B-C**). NARFL deletion resulted in decreased cytoplasmic cis-aconitase activity and intracellular glutathione and glutamine levels in endothelial cells, which were partially restored by Ferrostain-1 (**Figure 3D-F**).

Furthermore, Cell Rox kit was employed to assess the oxidative stress levels of endothelial cells. The findings demonstrated that NARFL deletion led to increased oxidative stress levels, and Ferrostain-1 partially alleviated the oxidative stress induced by NARFL gene deficiency (**Figure 4A-B**).

### NARFL Deletion Induces Endothelial Cell Dysfunction *in vitro*

Immunofluorescence analysis demonstrated that the deletion of *NARFL* gene resulted in down-regulation of CD31, a marker of endothelial cells, and up-regulation of α-SMA, a marker of fibroblasts^20^, consistent with the immunohistochemical features observed in the lung tissue of the proband (**Figure 4C**). We further investigated the impact of NARFL deletion on mitochondrial respiratory function and glycolysis rate using wild-type HPMECs, HPMECs with NARFL deletion, and HPMECs transfected with mutant *NARFL (c.482 G > T)* plasmid. The results revealed that mitochondrial respiratory function decreased in HPMECs with mutant *NARFL (c.482 G > T)* (**Figure 4D-F**), though the difference was minimal compared to wild-type HPMECs. However, NARFL deletion significantly reduced mitochondrial respiratory function without affecting glycolysis rate **(Figure 5A-C in the online-only Data Supplement**). Moreover, NARFL deletion led to decreased expression of CD31 in HPMECs, and narfl deletion in zebrafish resulted in decreased expression of cyp2p8 (CYP2JP in humans).

**Figure 5.**
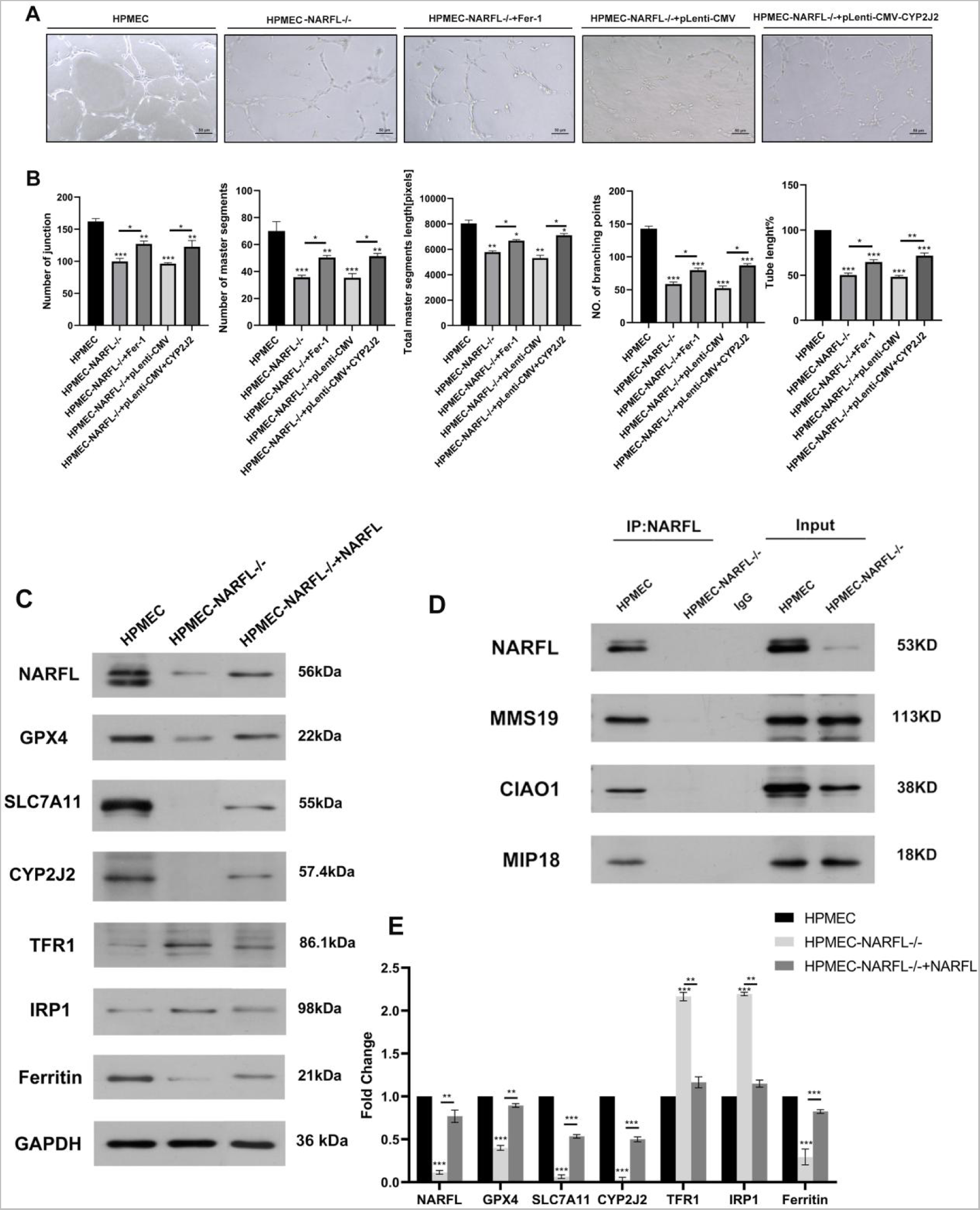
NARFL Gene Deletion Leads to Endothelial Cell Dysfunction with Abnormal Ferroptosis Pathway and Impaired Interaction with CIA System Related Proteins in Endothelial Cells. (**A-B**) Tube formation experiment comparing HPMEC, *NARFL-/-* HPMEC, *NARFL-/-* HPMEC treated with the Ferrostain-1 inhibitor of ferroptosis, and *NARFL-/-* HPMEC transfected with CYP2J2 plasmid. Results demonstrated the difficulty in tubular formation due to NARFL deletion, with partial recovery observed with Ferrostain-1 treatment and NARFL overexpression through CYP2J2 plasmid transfection. (**C**) Western blot analysis of HPMEC wild-type cells, *NARFL-/-* HPMEC cells, and *NARFL-/-* HPMEC cells transfected with an overexpression *NARFL* plasmid. The results showed downregulation of GPX4, SLC7A11, and Ferritin expression along with upregulation of TFR1 and IRP1 in NARFL-deficient cells. Additionally, CYP2J2 expression was negligible in NARFL-/- HPMEC, while transfection with the overexpressed *NARFL* plasmid partially restored these changes. (**D**) Co-immunoprecipitation analysis revealed that the *NARFL* mutant group failed to interact with MMS19, CIAO1, and MIP18 proteins, highlighting impaired interaction with CIA system-related proteins due to *NARFL* gene deletion.

To assess the impact of *NARFL* gene deletion on endothelial cell function, we compared tubule formation ability in wild-type HPMECs, NARFL-deleted HPMECs, NARFL-deleted HPMECs treated with Ferrostain-1, and NARFL-deleted HPMECs transfected with *CYP2J2* plasmid (**Figure 5A**). The results demonstrated that NARFL deletion impaired tubule formation, while treatment with Ferrostain-1 and overexpression of CYP2J2 partly restored this function (**Figure 5B**). Furthermore, the Evans Blue cell osmotic assay revealed that NARFL deletion led to impaired cell osmotic ability, which could be partially restored by Ferrostain-1, an ferroptosis inhibitor, and overexpression of *CYP2J2* plasmid (**Figure 5D-F in the online-only Data Supplement**).

### NARFL Deletion Results in Abnormal Ferroptosis Pathway and Disruption of CIA System-Related Protein Interaction in Endothelial Cells

Thus far, our findings suggest that NARFL deficiency leads to increased intracellular iron levels and oxidative stress, ultimately triggering ferroptosis. However, the specific underlying mechanism remains unclear. Intracellular iron metabolism begins with the binding of iron to transferrin in circulation, followed by endocytosis into cells through binding with the transferrin receptor 1 (TFR1) on the cell membrane, forming an unstable iron pool. Some of the iron is stored in the cytoplasm in the form of ferritin. In cells undergoing ferroptosis, the levels of iron and transferrin increase, while the amount of membrane iron transporter decreases^21^. Glutathione peroxidase 4 (GPX4) serves as a key regulator of ferroptosis^22^, converting glutathione (GSH) into oxidized glutathione to prevent cytotoxic lipid peroxidation and protect cells from ferroptosis. The GPX4 pathway is regulated by the cystine transporter system Xc^-^ (composed of catalytic subunit SLC7A11 and chaperone subunit SLC3A2). Cystine uptake mediated by SLC7A11 plays a crucial role in inhibiting oxidative reactions and maintaining cell survival under oxidative stress. Using Western blotting, we observed that NARFL deficiency led to down-regulated expression of GPX4, SLC7A11, and Ferritin, while TFR1 and IRP1 were up-regulated. In the zebrafish model study, NARFL deletion resulted in decreased expression of the iron-sulfur protein CYP2J2. In the HPMEC cell model, we found that NARFL deletion significantly reduced CYP2J2 expression, and transfection and overexpression of *NARFL* plasmid in NARFL deletion HPMECs partially restored the above changes. These results indicate that NARFL down-regulation not only up-regulates IRP1, which subsequently up-regulates TFR1 and down-regulates Ferritin, but also inhibits SLC7A11 and GPX4, activating the ferroptosis pathway (**Figure 5C, E**). NARFL is considered as the initiator of the cytoplasmic iron-sulfur protein assembly system known as the CIA system, which further transports mitochondrial iron-sulfur protein clusters. The CIA targeting complex (CTC), composed of CIAO1, MIP18, and MMS19, interacts with NARFL and facilitates the embedding of iron-sulfur clusters into specific apoproteins. Consistent with these findings, we observed that NARFL failed to bind to CIAO1, MIP18, and MMS19 in NARFL-deletion HPMECs, as determined by immunoprecipitation. Notably, NARFL appeared to bind with CIAO1 first, as down-regulation of NARFL directly resulted in decreased CIAO1 expression, while the expression of MMS19 and MIP18 remained unaffected. Thus, NARFL deletion leads to the disruption of normal transmission of mitochondrial iron-sulfur clusters to CIA system-related proteins and the failure of iron-sulfur clusters to embed into specific apoproteins, ultimately affecting the synthesis and maturation of cytoplasmic iron-sulfur proteins (**Figure 5D**).

### Deletion of Ciao3 Results in Embryonic Mortality and Impaired Vascular Development in Mice

In a study conducted by Song et al^11^., it was revealed that deletion of Ciao3 (NARFL homolog) led to embryonic lethality, with all Ciao3 knockout embryos being absorbed before 10.5 days of development. To further elucidate the underlying mechanism of lethality caused by Ciao3 knockout, embryos were collected at various time points including 8.5 days, 10.5 days, 12.5 days, and 13.5 days for genotype identification. The results demonstrated that Ciao3 knockout embryos persisted until 12.5 days, while complete absorption of Ciao3 knockout embryos occurred at 13.5 days and later stages (**Figure 6A**). Histological examination using hematoxylin and eosin (H&E) staining revealed significantly slower development and impaired vascular system development in Ciao3 knockout embryos compared to wild-type embryos. The yolk sac blood vessels in homozygous Ciao3 knockout mice displayed thinning, reduced branching, incomplete vascular network, decreased compactness, and blocked vascular development (**Figure 6B**). Based on these findings, we hypothesized that abnormal vascular development could be responsible for the lethality observed in Ciao3 knockout mice. During embryogenesis, endothelial cells play a critical role in cardiovascular system development, with these cells originating from blood islands formed from the mesoderm. To validate our hypothesis, immunofluorescence staining was performed to detect CD31 (**Figure 6C**) and CD34 (**Figure 6D**) markers of endothelial progenitor cells in whole embryos. The findings revealed disordered vascular structures, damaged and irregular vascular lumens in *Ciao3-/-* mouse embryos. In contrast, wild-type mouse embryos exhibited well-connected vascular networks. Additionally, the positive staining intensity of CD31 in endothelial cells and endothelial progenitor cells was notably reduced in *Ciao3-/-* embryos. These observations indicated maturation defects in vascular endothelial progenitor cells and endothelial progenitor cells in *Ciao3-/-* embryos. Based on the results obtained from zebrafish and cell models, it is speculated that embryonic lethality resulting from Ciao3 deletion may be attributed to increased oxidative stress and lipid peroxidation levels. To verify this hypothesis, the positive rates of 4-hydroxynonenal (4-HNE) (**Figure 6E**) and BODIPY (**Figure 6E**) were significantly higher in *Ciao3-/-* mouse embryos compared to wild-type embryos, as evidenced by fluorescence staining. Furthermore, using the γ-H2AX method, it was observed that DNA damage in *Ciao3-/-* mouse embryos was significantly augmented compared to wild-type embryos (**Figure 6H**).

**Figure 6.**
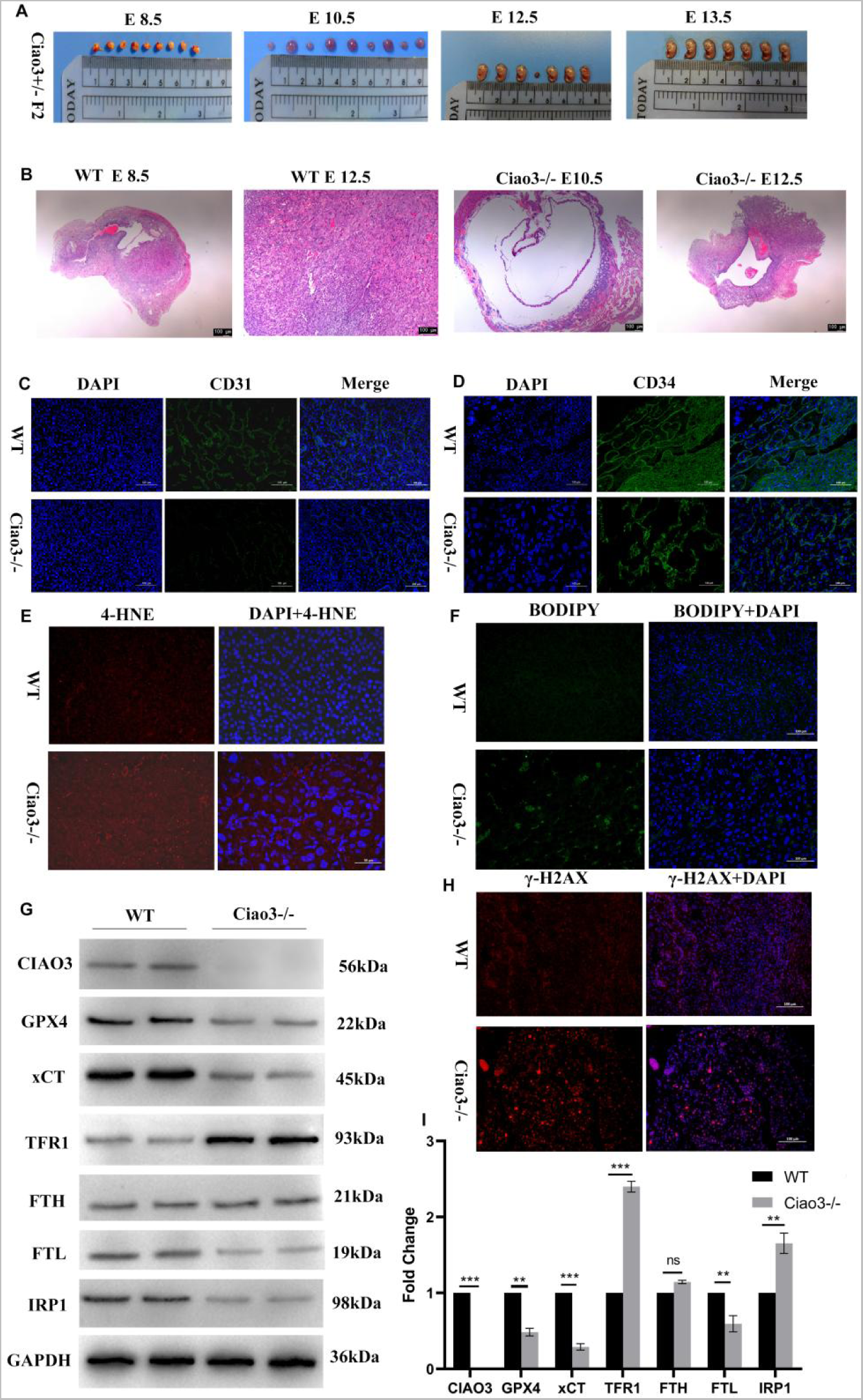
Deletion of Ciao3 Gene Leads to Embryonic Death and Vascular Development Disorder in Mice. (**A**) The morphological changes of Ciao3 heterozygous offspring at 8.5, 10.5, 12.5, and 13.5 days of gestation. Ciao3 knockout mice embryos were still present at 12.5 days but completely absorbed at 13.5 days and beyond. (**B**) H&E staining of wild-type and Ciao3-/- mice embryos at 8.5 and 12.5 days. The development of Ciao3 knockout embryos was significantly delayed compared to wild-type embryos, with a blockade in vascular system development. (**C**) Immunofluorescence staining of the endothelial marker CD31 in 12.5-day mice embryos. (D) Immunofluorescence staining of the endothelial progenitor cell marker CD34 in 12.5-day mice embryos. (**E-F**) Detection of 4-HNE (**E**) and BODIPY (**F**) in 12.5-day mice embryos. The positive rate of 4-HNE and BODIPY staining was significantly higher in Ciao3-/- embryos compared to wild-type embryos. (**G-I**) Western blot analysis showing significant downregulation of GPX4, xCT, and FTL, and significant upregulation of TFR1 and IRP1 in Ciao3 knockout embryos compared to wild-type embryos. (**H**) γ-H2AX detection showed enhanced DNA damage in Ciao3-/- embryos compared to wild-type embryos.

### Deletion of Ciao3 Results in Altered Expression of Ferroptosis Pathway-Related Proteins in Mouse Embryos

In our *in vitro* experimental cell model, we observed that down-regulation of NARFL resulted in decreased expression of GPX4, SLC7A11, and Ferritin, while the expression of TFR1 and IRP1 was upregulated. To investigate if a similar regulatory pathway exists in vivo, we performed Western blot experiments on 12.5-day-old wild-type embryos and Ciao3 knockout mouse embryos. The analysis revealed that the expressions of GPX4, xCT, and FTL were down-regulated, whereas TFR1 and IRP1 were upregulated in Ciao3 knockout mouse embryos. These findings align with the results obtained from the cell model, suggesting that the deletion of Ciao3 leads to the upregulation of IRP1 expression in the mice model. This, in turn, drives the upregulation of TFR1 expression and the downregulation of FTL expression. Ciao3 deletion also inhibits the downregulation of xCT and GPX4 expression, resulting in the activation of the ferroptosis pathway (**Figure 6G, I**).

### Impairment of Vascular Function in Ciao3 Heterozygous Mice

Although *Ciao3+/-* mice show minimal differences in appearance compared to wild-type mice, their activity levels visibly decrease from the 8th to 9th week of age. *Ciao3+/-* knockout mice also exhibit reduced activity compared to wild-type mice. At the 9th week, the heart, lung, and liver of mice from both groups were dissected and stained with H&E. No differences were observed in the heart and liver; however, significant differences were observed in the pulmonary vessels (**Figure 7A**). The lungs of *Ciao3+/-* mice were significantly thicker compared to those of wild-type mice. Immunohistochemical staining of endothelial cell marker CD31 showed even distribution and dense expression in the lung lobes of wild-type mice, while α-smooth muscle actin (α-SMA) expression was minimal (**Figure 7B**). In contrast, CD31 expression was significantly reduced, and α-SMA expression was increased, in the lung lobes of *Ciao3+/-* mice. These findings suggest a decrease in endothelial cells and an increase in smooth muscle cells in the lungs of *Ciao3+/-* mice, which may contribute to thickening of the pulmonary artery wall and stenosis of the pulmonary artery lumen. To investigate whether vascular endothelial cell function was compromised in *Ciao3+/-* mice, angiogenesis and vascular permeation experiments were conducted. Aortic rings from mice were cultured in an extracellular matrix (ECM) medium for 4 days. The results revealed a significant reduction in the number of buds in the aortic rings of *Ciao3+/-* mice compared to wild-type mice (**Figure 7C-D**), indicative of inhibited angiogenesis in *Ciao3+/-* mice. Furthermore, vascular permeability assays using albumin-bound Evans Blue stain solution showed significantly darker staining in the aortic arch and cerebral vessels of *Ciao3+/-* mice compared to the control group, suggesting increased vascular permeability in *Ciao3+/-* mice. These results collectively indicate impaired vascular function in Ciao3-deficient mice.

**Figure 7.**
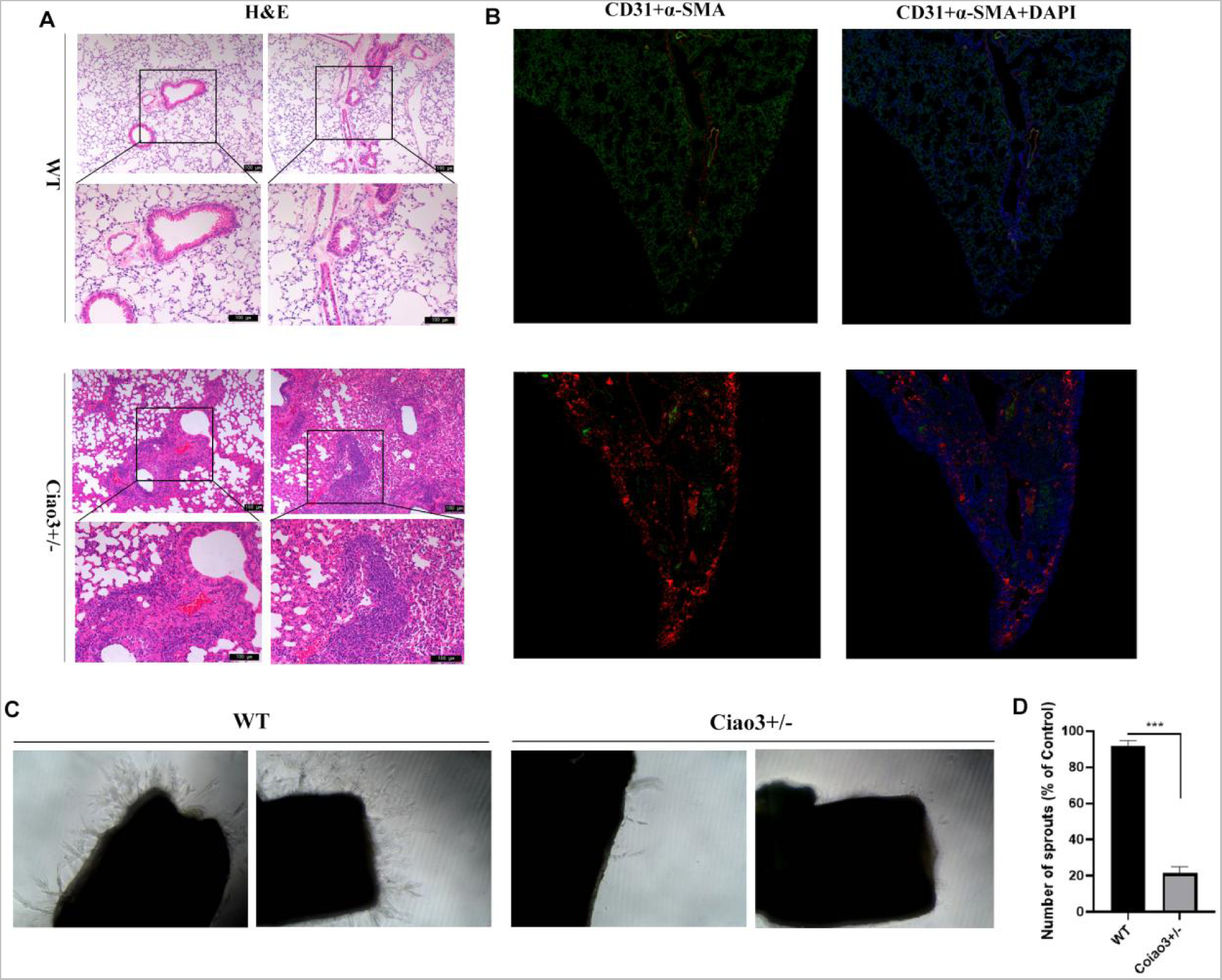
Impairment of Vascular Function in Ciao3 Heterozygous Mice. (**A**) H&E staining of lung sections from 9-week-old mice. The pulmonary artery walls and small blood vessels were significantly thicker in Ciao3+/- mice compared to wild-type mice. (**B**) Dual fluorescence immunostaining of CD31 and α-SMA in lung sections of 9-week-old mice. CD31 expression was evenly distributed in the lung lobes of wild-type mice, with dense CD31-positive cells, while α-SMA expression was sparsely distributed. (**C-D**) Aortic rings embedded in matrix glue were cultured in ECM medium for 4 days. The number of sprouts in the aortic rings of Ciao3+/- mice was significantly reduced compared to wild-type mice.

### *NARFL* Polymorphisms are Susceptible Sites for Vascular Endothelial Dysfunction Diseases

A total of 387 cases of vascular endothelial dysfunction and 409 control individuals were included in this study. The control group consisted of 409 healthy individuals with an average age of 48 ± 12.5 years, including 228 males and 181 females. The case group had a mean age of 50 ± 17.2 years, including 216 males and 171 females. There were no significant differences in age and sex distribution between the case and control groups. The basic characteristics of the 387 cases are provided in **Table S1**, where 20.4% were drinkers and 79.6% were non-drinkers. In addition, 38.2% were smokers and 61.8% were non-smokers. Among the cases, 187 patients had pulmonary hypertension with obvious pulmonary vein or pulmonary capillary involvement, 51 patients had neurodegenerative diseases, 39 patients had epilepsy, 66 patients had systemic lupus erythematosus, and 44 patients had rheumatoid arthritis and arteritis. Echocardiography revealed mild reflux in 167 cases (43.2%), moderate reflux in 75 cases (19.4%), and severe reflux in 10 cases (2.5%).

The study of NARFL originated from a rare family with pulmonary arterial hypertension secondary to diffuse pulmonary arteriovenous malformation, where a missense mutation on NARFL caused severe consequences. Typically, in genetic studies, we identify the pathogenic mutation of a gene through the phenotypic presentation in a family, and then explore the underlying pathogenic mechanism. Studying rare diseases and their pathogenesis is both challenging and meaningful. However, based on the phenotype observations of Ciao3 hybrid mice at later stages, we questioned whether there are susceptible sites of *NARFL* gene polymorphisms that contribute to vascular endothelial dysfunction diseases. Although these susceptible sites may not cause severe phenotypes in rare PAH disease, they have the potential to increase susceptibility to endothelial dysfunction diseases.

Among the seven tagSNPs of *NARFL* (rs61112891, rs2071952, rs117952680, rs9928077, rs3752556, rs11248948, and chr16-731143), the distribution of rs1179252680, rs2071952, rs61112891, and chr16-731143 showed statistical differences between cases and controls (**Figure 8A, able S2**). The odds ratio (OR) values of rs1179252680, rs2071952, and rs61112891 were greater than 1, indicating that these variants may act as risk factors for vascular endothelial dysfunction, while chr16-731143 had an OR value less than 1, indicating its potential role as a protective factor. Our genotype analysis revealed that individuals carrying the GG and CG genotypes of rs61112891 had a significantly increased risk for vascular endothelial dysfunction, with the OR of the GG genotype being 3.971 higher than the CG genotype (1.328). Moreover, carriers of the CT and TT genotypes of rs2071952 had a significantly increased risk for vascular endothelial dysfunction compared to controls, with no individuals in the control group found to have the TT genotype. Additionally, individuals carrying the GA genotype of rs117952680 had a significantly increased risk, with an OR value of 5.284. Furthermore, carriers of the GG genotype of rs11248948 had a significantly increased risk of vascular endothelial dysfunction, with no individuals in the control group found to have the GG genotype. Finally, carriers of the TC or CC genotypes of chr16-731143 had a reduced risk of vascular endothelial dysfunction, with an OR value of 0.69. No differences in the genotype distributions of rs9928077 and rs3752556 were observed between the two groups.We further conducted a genotype frequency distribution analysis for seven tagSNPs in patients with various vascular endothelial dysfunction-related diseases, including pulmonary hypertension, neurodegenerative diseases, epilepsy, systemic lupus erythematosus, rheumatoid arthritis, and arteritis (**Figure 8B, Table S3**). Our results indicated that carrying the rs11248948 (GG) genotype significantly increased the risk of cerebral small vessel epilepsy (OR=3.588, *p*=1.293×10^-12^). Additionally, carrying the rs117952680 (GA) genotype significantly increased the risk of cerebral small vessel epilepsy (OR=5.826, *p*=0.005), neurodegenerative diseases (OR=7.129, *p*=1.62×10^-4^), and pulmonary hypertension patients with obvious pulmonary vein or pulmonary capillary involvement (OR=6.318, *p*=1.175×10^-7^). Moreover, carrying the rs2071952 (TT or CT) genotype significantly increased the risk of cerebral small vascular epilepsy (OR=3.462, *p*=2.176×10^-10^), while carrying the rs611289 (GG or CG) genotype significantly increased the risk of cerebral vascular epilepsy (OR=2.699, *p*=3.982×10^-8^). On the other hand, carrying the chr16-731143 (TC or CC) genotype significantly reduced the risk of pulmonary hypertension (OR=0.669, *p*=0.002), systemic lupus erythematosus (OR=0.546, *p*=0.005), cerebral small vessel epilepsy (OR=0.489, *p*=0.015), and rheumatoid arthritis and arteritis (OR=0.433, *p*=0.003) in patients with obvious pulmonary vein or pulmonary capillary involvement. No significant differences were observed between the genotypes of other tagSNPs and the analyzed diseases. Furthermore, we analyzed the expression of NARFL in both the case and control groups, revealing significantly lower NARFL expression levels in the case group compared to the control group (**Figure 6B in the online-only Data Supplement**). Furthermore, a negative correlation was observed between NARFL expression levels and MDA, a biomarker associated with ferroptosis (**Figure 6C in the online-only Data Supplement**). The receiver operator characteristic (ROC) curve analysis showed that NARFL expression level was more effective in differentiating tagSNP groups, with an area under the curve (AUC) of 0.765, compared to MDA (AUC=0.540), indicating a lower discriminatory power for tagSNP groups (**Figure 6D in the online-only Data Supplement**). Collectively, these findings suggest that decreased NARFL expression may be associated with an increased risk of vascular endothelial dysfunction in individuals with specific tagSNPs.

**Figure 8.**
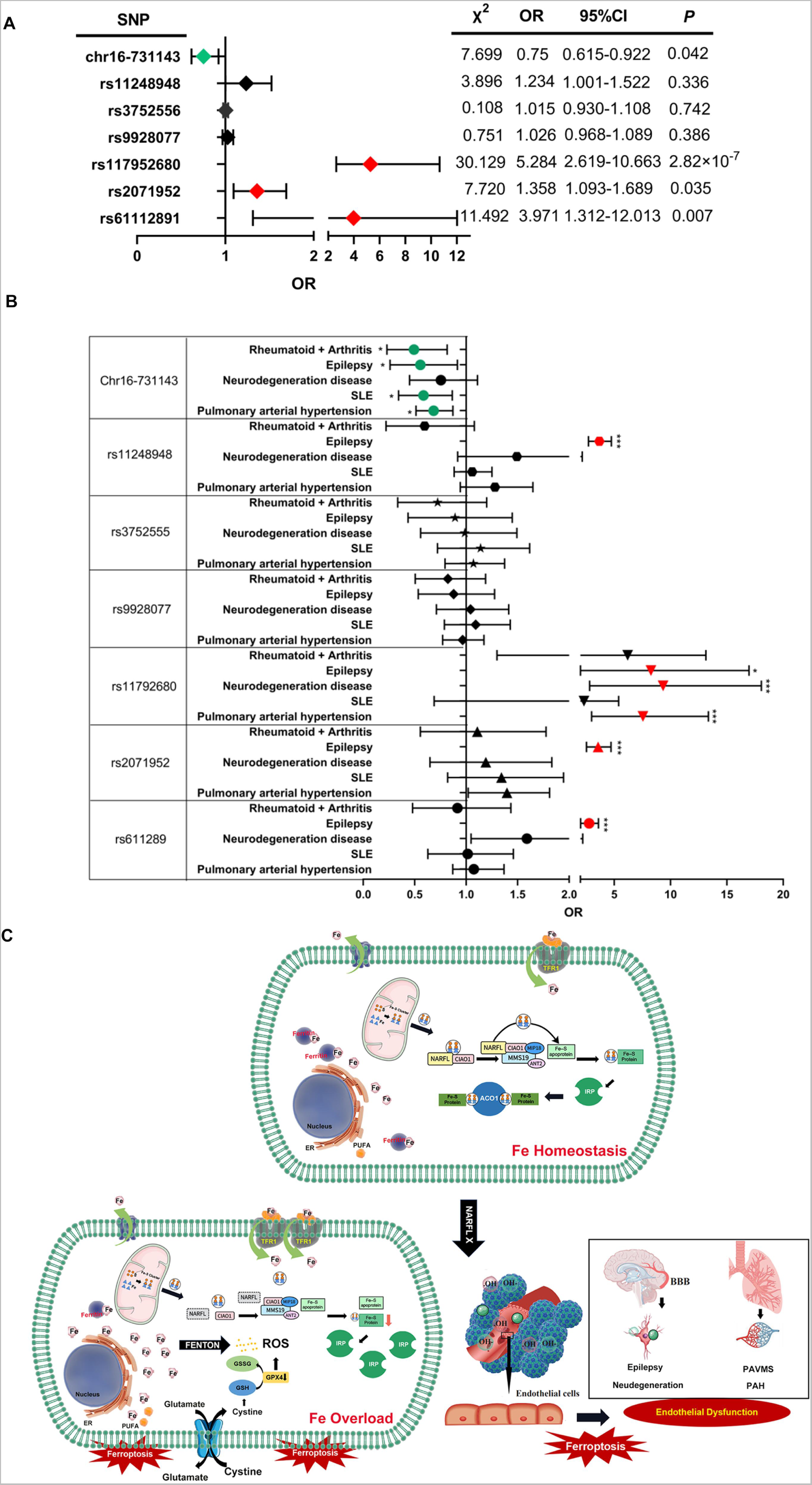
NARFL Polymorphisms as Susceptible Sites of Vascular Endothelial Dysfunction Diseases. (**A**) Among the seven tagSNPs of NARFL (rs61112891, rs2071952, rs117952680, rs9928077, rs3752556, rs11248948, and chr-731143), the frequency distribution of four SNP genotypes (rs1179252680, rs2071952, rs61112891, and chr16-731143) showed statistically significant differences between cases and controls. The genotypes of rs1179252680 (*p* < 0.001), rs61112891 (*p* < 0.01), rs2071952 (*p* < 0.05), and chr16-731143 (*p* < 0.05). The odds ratio (OR) values of rs1179252680, rs2071952, and rs61112891 were greater than 1, while the OR values of chr16-731143 were less than 1. (**B**) The genotype frequency distribution was further analyzed between the seven tagSNPs and patients with various vascular endothelial dysfunction-related diseases, including pulmonary hypertension, neurodegenerative diseases, epilepsy, systemic lupus erythematosus, rheumatoid arthritis, and arteritis. (**C**) Mechanism Summary Diagram: under normal circumstances, NARFL facilitates the transfer of iron-sulfur clusters (ISC) synthesized in mitochondria to the cytosolic iron chaperone (CTC), composed of CIAO1, MIP18, and MMS19, through the interaction with CIAO1. This transfer of ISC allows for the incorporation of the ISC into specific apo-proteins, forming mature ferritin and maintaining intracellular iron homeostasis. However, in the absence of NARFL, the transfer of ISC from mitochondria to CTC is hindered, leading to the failure in the formation of mature ferritin. Consequently, iron-responsive protein 1 (IRP1) increases the expression of transferrin receptor 1 (TFR1), promoting iron uptake, while inhibiting the expression of ferritin, inhibiting iron storage. As a result, intracellular iron levels increase, triggering enhanced oxidative stress through the Fenton reaction. Concurrently, the downregulation of SCL7A11 and GPX4 reduces the synthesis of cytoplasmic glutathione and glutamine, exacerbating reactive oxygen species production. This oxidative stress activates lipid peroxidation, inducing ferroptosis and vascular endothelial cell death, ultimately leading to vascular endothelial dysfunction.

## DISCUSSION

These findings demonstrate that *NARFL* gene knockout leads to endothelial dysfunction and further abnormal vascular development in HPMECs, zebrafish and mice. This is characterized by a weak endothelial structure and decreased tube formation ability, which are the developmental structural basis for the death of homozygous gene knockout zebrafish and mouse embryos. In HPMECs cell model studies, it was found that the deletion of the NARFL gene prevents the transmission of mitochondrial synthesized ISC to CTC, resulting in the inability to form mature iron-sulfur proteins. ACO1, which acts as a representative of iron-sulfur protein, becomes an “RNA binding protein” IRP1 when cytoplasmic ACO1 cannot obtain ISC. IRP1 increases the expression of TFR1 and inhibits the expression of Ferritin, resulting in increased iron intake. This leads to increased intracellular iron ions, enhanced oxidative stress, and down-regulation of SCL7A11 and GPX4. The decrease in cytoplasmic glutathione and glutamine synthesis further increases the production of reactive oxygen species, activates lipid peroxidation, and induces vascular endothelial cell death and dysfunction. The *NARFL* gene polymorphisms rs11248948 (GG type), rs117952680 (GA type), rs2071952 (TT or CT type), and rs611289 (GG or CG) significantly increase the risk of cerebral small-vessel epilepsy, degenerative disease, and pulmonary hypertension, respectively (**Figure 8C**).

In the zebrafish model^17^ of *NARFL* knockout and combined with Tg (*flk*: eGFP) model which is suitable for studying vascular development and morphology, it was found that *NARFL* gene knockout not only leads to death of juvenile fish but also epilepsy-like abnormal behavior, abnormal blood-brain barrier (BBB) morphology, and neuronal lesions. In Liu’s study, NARFL mutations were also found in one epileptic family^23^.The pathogenesis of epilepsy is complex, and one of the mechanisms is the imbalance of central nervous system homeostasis caused by BBB injury^15,24^. Endothelial cells in the vascular barrier contain numerous ATP-binding cassette transporters (ABC transporters) that help maintain central nervous system homeostasis and prevent the passage of harmful substances through the BBB^16^. Furthermore, our research revealed that NARFL deletion led to abnormal vascular development and structure in zebrafish. Previous studies have already shown that knocking out NARFL resulted in abnormal intestinal vessels in zebrafish. To better understand the process of vascular development in zebrafish, we used the *Tg (flk: eGFP)* model and observed that NARFL deletion caused deformity or even absence of dorsal longitudinal anastomosis vessels and connecting cells, as well as distortion of the dorsal aorta and PCV. Subsequent experiments demonstrated that NARFL deletion led to increased oxidative stress, lipid peroxidation, and iron levels. It is speculated that the increase in lipid peroxidation, caused by the rise in free iron, along with the significant increase in oxidative stress, leads to injury of vascular endothelial cells in zebrafish and results in blood vessel malformation during development. When measuring the blood flow of zebrafish, we discovered that the blood flow of *narfl-/-* zebrafish significantly increased before 6 dpf, reached its peak at 6 dpf, and then decreased significantly after 6 dpf. However, the blood flow velocity of wild zebrafish did not show significant changes. To investigate the underlying reasons, we examined the functional markers ET-1 and NO in endothelial cells and found that ET-1, responsible for vasoconstriction, increased significantly at 6 dpf, while NO, responsible for vasorelaxation, decreased significantly at 6 dpf. This phenomenon, however, did not occur at 4 dpf. We hypothesize that endothelial cells compensate for the injury before 6 dpf, but at 6 dpf, the relaxation and contraction functions of endothelial cells are significantly impaired. Almost all *narfl-/-* zebrafish died before 13 dpf, indicating that narfl deficiency seriously damages the function of vascular endothelial cells. Furthermore, our observations revealed that the dorsal aorta and posterior aorta of *narfl-/-* zebrafish did not fuse to form a regular circular circulation, as seen in wild-type zebrafish. Instead, they formed a distorted and disorganized shape. The formation of the dorsal aorta and PCV occurs during the early embryonic angiogenesis stage. When zebrafish reaches approximately 15 nodules, angioblasts in the middle layer of the lateral plate converge at the midline to form the dorsal aorta and PCV^25^. However, in *narfl-/-* zebrafish, abnormalities occur during the process of angiogenesis, generation, and differentiation, disrupting the normal development of the vascular network.

Transcriptome sequencing revealed that cyp2p8 in zebrafish (CYP2J2 in humans) is an important cytochrome P450 monooxygenase involved in the metabolism of polyunsaturated fatty acids (PUFA) in the cardiovascular system^26, 27^. The mechanism of action for cyp2p8 involves using molecular oxygen to insert an oxygen atom into the substrate and reducing the second oxygen atom into water molecules. NADPH, provided by cytochrome P450 reductase (NADPH-cytochrome P450 reductase), supplies two electrons necessary for the epoxidation of PUFA double bonds. This conversion leads to the formation of four regionally isomeric Epoxyeicosatrienoic acids (EETs), which may play a crucial role in the epoxidation of endogenous cardiac arachidonic acid pools. CYP2J2 is widely expressed in vascular endothelial cells^28^. It and its products have been found to exert protective effects on vascular injury. CYP2J2 can convert hydrogen peroxide into hydroxyepoxy metabolites and participate in eicosanoic acid metabolism. It can also interact with 15-lipoxygenase to metabolize arachidonic acid and convert hydroperoxicosatetraenoates (HpETEs) into hydroxy epoxy eicosatrienoates (HEETs)^29^. HEETs have been shown to play a protective role in vascular injury through various mechanisms, including anti-inflammation, anti-apoptosis, and inhibition of vascular endothelial cell aging^29, 30^. In summary, a decrease in the expression of CYP2J2 results in a reduction in HEET levels, thereby diminishing the protective effect on blood vessels. Through qRT-PCR and WISH experiments, it was found that cyp2p8 (CYP2J2 in humans) is expressed in the blood vessels of wild zebrafish. However, the expression of cyp2p8 in NARFL-/- zebrafish blood vessels was significantly decreased, suggesting a potential link between NARFL deficiency and impaired metabolism of PUFA in the cardiovascular system. Further research is required to fully understand the role of NARFL and its interaction with cyp2p8/CYP2J2 in vascular development and homeostasis.

In the preface, it is mentioned that the deletion of NARFL hinders the transmission of ISC to the IRP1 protein. As a result, ACO1 loses ISC and becomes IRP1, leading to a decrease in ACO1 activity. Previous studies have reported similar phenotypes when mitochondrial ISC synthesis-related proteins are deleted in both lower yeast and higher mammalian cells^31–34^. These phenotypes include excessive iron in mitochondria, increased oxidative stress, blocked electron transmission, and decreased mitochondrial function. Additionally, a decrease in cytoplasmic iron levels up-regulates IRP1 activity, resulting in increased iron uptake. Our research found that the deletion of NARFL does not affect mitochondrial cis-aconitase activity but decreases cytoplasmic cis-aconitase activity. This leads to iron overload, which prompted us to investigate whether mitochondrial function is affected by NARFL. By using the seahorse XFe analyzer, we observed a decrease in mitochondrial respiratory function due to NARFL deletion. We speculate that this mechanism may be attributed to the increased activity of IRP1 caused by NARFL deletion. This leads to the up-regulation of transferrin receptor (TFR) expression, resulting in increased intracellular iron uptake. However, the iron obtained from mitochondria cannot be effectively utilized, leading to aggravated iron overload, increased oxidative stress, and hindered electron transfer within mitochondria.To verify this mechanism, further cell experiments are needed.

Corbin et al. conducted a genome-wide DNA and RNA array analysis combined with functional genomics research and discovered that *NARFL* gene overexpression was present in two oxygen-resistant strains of HeLa cells. They also found that hyperoxia-induced overexpression of NARFL can protect the activity of iron-sulfur proteins, specifically ACO1, highlighting the crucial role of *NARFL* in resisting oxidative stress caused by hyperoxia^35–37^. Furthermore, similar to the results obtained from knocking out the NARFL homologue Nar1 in yeast, it was found that NARFL deletion leads to defects in cytoplasmic iron-sulfur protein assembly, ultimately resulting in cellular and organismal death. This further confirms the significant role of NARFL in the cytoplasmic iron-sulfur protein assembly pathway^38^. In a study by Fan XR et al^38^ it was discovered that the NARFL-S161I mutant was unable to bind to the functional CIA complex. It was speculated that this mechanism could be related to the development of diffuse pulmonary arteriovenous malformation, a condition associated with this specific mutation, which was first identified by the researchers. This study also revealed that the association between NARFL and the CIA complex is closely linked to cellular iron levels. The binding of NARFL to the CIA complex was found to be influenced by oxidative stress levels and hypoxia. Specifically, the interaction between NARFL and the CIA complex was enhanced when iron supplementation or hypoxic conditions were introduced, while the presence of reactive oxygen species weakened the interaction between NARFL and the components of the CIA complex.

According to the results of experiments on zebrafish and cell models, it can be concluded that NARFL plays a crucial role in maintaining iron homeostasis in cells. Under normal conditions, NARFL facilitates the transfer of iron-sulfur clusters (ISC) synthesized in mitochondria to the cytoplasmic iron-sulfur protein assembly complex (CTC). The CTC is composed of CIAO1, MIP18, and MMS19 and interacts with CIAO1 to form mature iron-sulfur proteins and maintain iron balance in cells. However, in the absence of NARFL, ISC cannot effectively transfer to the CTC, resulting in the inability to form mature iron-sulfur proteins. One example is ACO1, an iron and sulfur representative that acts as both an enzyme and an “RNA binding protein” IRP1. Without ISC, cytoplasmic ACO1 cannot function properly and instead acts as IRP1. IRP1 then increases the expression of transferrin receptor 1 (TFR1) and inhibits the expression of ferritin, leading to increased iron uptake in cells. The decrease in ferritin levels prevents the normal binding and storage of iron, resulting in an increase in intracellular iron ions and significant oxidative stress through the Fenton reaction.The down-regulation of SCL7A11, GPX4, and CYP2J2, along with the decrease in glutathione and glutamine synthesis in the cytoplasm, further enhances the production of reactive oxygen species (ROS) and activates lipid peroxidation. This ultimately leads to ferroptosis-induced vascular endothelial cell death and dysfunction. ACO1 serves as a crucial link between iron metabolism balance and the oxidative stress signaling pathway. The decrease in ACO1 activity and the increase in free iron content in the cytoplasm promote ROS production.In the cytoplasm, IRP1 acts as an iron receptor. When cellular iron levels increase, IRP1 binds to [4Fe-4S] clusters and converts into ACO1. Conversely, when cellular iron levels decrease, IRP1 dissociates from [4Fe-4S] clusters and binds to iron-responsive elements (IREs) in the non-coding region of iron metabolism-related protein mRNA. This binding promotes iron absorption and reduces iron storage in cells, thereby restoring cellular iron levels. Notably, the mRNA of transferrin receptor (TFR) contains five IRE structures in its 3’UTR. When iron-deficient, IRP can bind to these IREs and protect TFR mRNA from degradation, leading to an increase in TFR levels and iron absorption in cells. The IRE of ferritin is present in its 5’UTR, and when iron-deficient, IRP1 binds to the ferritin IRE, reducing ferritin synthesis and resulting in decreased iron storage and utilization in cells^39^. *NARFL* plays a critical role in maintaining this delicate balance. Once NARFL is deleted, this balance is disrupted, leading to dysregulation of iron metabolism and oxidative stress in cells.

Song et al. is the only team that has completed the research report of mice Ciao3 gene knockout^11^. The team discovered that mice embryos died 10.5 days after the knockout of the Ciao3 gene. Additionally, inducing acute knockout of Ciao3 in adult mice resulted in their death, along with a significant decrease in cytoplasmic aconitase activity in their liver. Knockout of Ciao3 in mice embryonic fibroblasts led to a decrease in cytoplasmic aconitase activity and cell viability^11^. We observed that Ciao3-/- mice died or were absorbed in the early embryonic stage (E 12.5 and before), which aligns with Song’s conclusion that Ciao3-/- mice embryos died or were absorbed before 10.5 days. As the occurrence of blood vessels is one of the earliest events in embryonic development, with mesodermal cells differentiating into vascular cells such as hematopoietic progenitor cells and endothelial progenitor cells from the 7th day, we speculate that Ciao3 deletion could damage endothelial progenitor cells. Our findings indicate that Ciao3-/- mice embryonic endothelial progenitor cells indeed exhibited maturation defects, resulting in a failure to connect endothelial cells into a vascular network in the early embryo. Previous reports have shown that vascular endothelial progenitor cells form a functional circulation in the early stage^40^. Vascular progenitor cells respond to basic fibroblast growth factor and bone morphogenetic protein 4 in the posterior primitive stripe as flk1-positive mesodermal cells, which produce blood and endothelial cells simultaneously. However, after migrating to the outer and inner embryos, they are limited to either hematopoiesis or angiogenesis^41^. In the yolk sac, these endothelial progenitor cells aggregate into endothelial-lined blood islands, which then fuse to form a primary capillary plexus. This plexus undergoes remodeling with intracellular blood vessels to form a mature circulation. If the maturation cycle of endothelial progenitor cell formation is not established, the embryo ceases to develop and dies^42, 43^.

Compared to wild-type embryos, we observed a significant increase in oxidative stress levels, lipid peroxidation levels, and DNA double-strand breaks in Ciao3-/- mouse embryos. Excessive reactive oxygen species (ROS) during embryonic development can cause damage to DNA, proteins, and lipids, leading to mitochondrial damage. Mitochondrial DNA is essential for oxidative phosphorylation, and defects in embryonic mitochondrial DNA can result in metabolic dysfunction, embryo damage, developmental retardation, and even developmental stagnation^44^.The production of ROS is influenced by various factors, and the amount of ROS produced varies during different stages of embryo development. In mice embryos, ROS production is highest during fertilization and the G2/M stage of the second cell division, and iron ions can directly act on lipids and amplify the peroxidation damage caused by free hydroxyl radicals^45^.ipids play a crucial role in constituting the cytoskeleton, and lipid peroxidation occurs when polyunsaturated fatty acids combine with oxygen free radicals in vivo. Excessive oxidative stress leads to an increase in lipid peroxidation, which can dissolve polyunsaturated fatty acids in the cell membrane, disrupt cell membrane structure, alter cell membrane fluidity and permeability, and affect the transfer of cell metabolites and cell signal transduction^46^. Embryonic developmental stagnation is a self-protection mechanism to prevent abnormal or low-quality embryos from continuing to develop.Western blot experiments demonstrated that GPX4, xCT, and FTL expressions were down-regulated in Ciao3-/- mouse embryos, while the expression of TFR1 and IRP1 was up-regulated, consistent with the results of the cell model. Therefore, we infer that Ciao3 deletion leads to the up-regulation of IRP1 expression in the mouse model, which further up-regulates TFR1 expression and down-regulates FTL expression. Ciao3 deletion also induces the down-regulation of xCT and GPX4 expression, activating the ferroptosis pathway, increasing oxidative stress levels and lipid peroxidation levels in Ciao3-/- embryo endothelial progenitor cells, obstructing the maturation and circulation of endothelial progenitor cells, and ultimately resulting in early embryo death.We observed that the surface of Ciao3+/- mice did not differ significantly from that of wild-type mice, but we did observe obvious endothelial cell injury in the lungs of Ciao3+/- mice in the later stage. There was evident thickening of the wall of pulmonary blood vessels, a decrease in endothelial cells, and an increase in smooth muscle. In the mouse model, we also found that the absence of Ciao3 leads to impaired maturation and differentiation of endothelial progenitor cells. Endothelial progenitor cells differentiate into veins and arteries and gather in primitive capillaries. Neovascularization initially consists of endothelial cells. Vascular maturation requires the interaction of vascular and arterial factors for a sufficient duration, allowing endothelial cells to be tightened and covered by parietal cells and extracellular matrix. If endothelial cells are damaged, blood vessels may leak, become fragile, easily rupture and bleed, leading to reduced blood flow and vascular degeneration^47–49^.

Gene Curation^50^ is a process that involves extracting information from literature and databases to evaluate the strength of the “gene-disease” relationship based on current research. According to the ClinGen Gene Curation Standardized Evidence SOP version 8, NARFL has been reported in relation to pulmonary hypertension secondary to diffuse pulmonary arteriovenous malformation. Based on this evidence, NARFL is strongly associated with this condition.When the *NARFL* gene is knocked out, it can induce ferroptosis and increase oxidative stress levels, leading to endothelial cell injury. Additionally, certain susceptible sites in the *NARFL* gene polymorphism can increase the risk of vascular endothelial dysfunction diseases. Our findings indicate that carrying specific tagSNPs in the non-coding region of *NARFL* can significantly increase the risk of cerebral small vascular epilepsy, cerebral vascular epilepsy, degenerative diseases, and pulmonary hypertension with pulmonary vein or pulmonary capillary involvement. Although these tagSNPs are located in the non-coding region of NARFL, our later study revealed that the expression of NARFL was significantly lower in the case group compared to the control group. The expression levels of NARFL were more effective in predicting the presence of dangerous tagSNPs in the case group compared to MDA. However, it is still unclear whether these tagSNPs directly reduce the expression level of NARFL, and further evidence is needed to confirm this. The diseases most strongly associated with NARFL gene tagSNPs include cerebral small vessel epilepsy, degenerative diseases, and pulmonary hypertension with pulmonary vein or pulmonary capillary involvement. This is consistent with our previous findings in zebrafish and mouse models. Due to the limited number of specimens in this study, it is necessary to collect more samples to verify the associations between specific tagSNPs and the mentioned diseases. Due to the constrained number of specimens obtained in this research, there is a pressing need to gather additional samples. This will enable a more robust validation of whether individuals carrying the rs11248948 (GG), rs2071952 (TT), and rs611289 (GG) polymorphisms are at heightened risk for cerebellar vascular epilepsy. Furthermore, it’s imperative to investigate whether carriers of the rs117952680 (GA) variant are more susceptible to cerebellar vascular epilepsy and neurodegenerative diseases, as well as to pulmonary hypertension characterized by pronounced pulmonary vein or capillary involvement.

## Data Availability

All data referred to in this manuscript are available upon request

https://pan.baidu.com/s/1JIAKnpBzIsiwOMOYqosnAw?pwd=5f4w

## Authors

Hui Hu, MD, PhD; Jing Luo, MD, PhD; Li Yu, MD; Daoxi Qi, MD; Boyu Li, MD; Yating Chen, MD; Xiaokang Zhang, MD; Chen Wang, MD, PhD; Fan Wang, MD; Zhan Yin, PhD; Fang Zheng, MD, PhD.

## Correspondence

Fang Zheng, MD, PhD,Center for Gene Diagnosis, Zhongnan Hospital of Wuhan University, Wuhan, Hubei, 430071, China.

Zhan Yin, PhD, State Key Laboratory of Freshwater Ecology and Biotechnology, Institute of Hydrobiology, Chinese Academy of Sciences, Wuhan, Hubei, 430072, China.

## SUPPLEMENTAL MATERIAL

### Supplemental Methods

#### Zebrafish and behavioral analysis

The zebrafish culture and experimental procedures were conducted in accordance with the guidelines specified in the zebrafish handbook and regulations on the Care and use of Laboratory Animals. These protocols were approved by the Institute of Hydrobiology, Chinese Academy of Sciences (Approval ID: IHB 2013724). The wild type zebrafish used in the study were maintained under standard conditions1. Zebrafish developmental stages were determined based on hour post-fertilization (hpf) or days post-fertilization (dpf)^2^.

Homozygous *flk:GFP/narfl-/-* zebrafish lines were generated through mating of *flk:GFP/narfl+/-* adult fish, obtained by hybridizing *flk:GFP/Con* and *narfl+/-* fish. For behavioral analysis, 7 dpf zebrafish were individually placed in a 24-hole plate. The plate was then positioned in a behavior analysis system equipped with Viewpoint zebrafish tracking software (ViewPoint Life Sciences, Zebraoo1, Lyon, France). The detection area, along with parameters including time (60 min), background pixels (18-24 pixels), speed (0.4-10 mm/s), output interval time (60 s), and photocyte intensity (500 lx), were adjusted.

To ensure sufficient data for zebrafish of different genotypes, a total of 10 groups of experiments were conducted. The behavior of 240 zebrafish was monitored, and their track maps were recorded and analyzed using ViewPoint’s Micro Zebra Lab. This software utilizes high-speed video recordings of zebrafish to measure various parameters such as pulse rate, blood flow, and changes in vessel diameter. The blood flow data were calculated using the software algorithm, which analyzes the correlation between consecutive frames. Zebrafish aged 3-13 dpf were placed on a microscope slide for observation. The video file of the zebrafish sample was opened on a computer, and the microscope slide with the zebrafish was positioned in the designated observation area. A 1-minute analysis was conducted once the settings were verified. The original measured data is represented in red, while the results obtained after filtering with the fast Fourier Transform algorithm (FFT) are represented in blue.

#### Blood vessels and blood-brain barrier imaging in zebrafish

Confocal microscopy images of 3dpf *flk:GFP/narfl-/-* homozygous embryos were acquired using a Zeiss ISM 710 confocal microscope. The images captured the blood-brain barrier (BBB), Dorsal longitudinal aorta vessels (DLAV), Dorsal aorta (DA), Posterior cardinal vein (PCV), connector cells, and basal cells. The percentage of total DLAV, junction cells, and basal cells was calculated as previously described^3^.

The ultrastructure of the blood-brain barrier in zebrafish was observed using transmission electron microscopy. 7 dpf zebrafish with different genotypes were fixed, dehydrated, made transparent, embedded in wax, sectioned (thickness of 6 μm), stained with toluidine blue, and examined for Nissl bodies under a microscope. Additionally, a Prussian blue staining solution was prepared by mixing hydrochloric acid and potassium ferricyanide in a 1:1 ratio. Prussian blue staining was performed using the same method mentioned above to observe the presence of positively stained Prussian blue complexes.

#### Whole-mount RNA in situ hybridization (WISH) and quantitative real time PCR

The primers used were as follows: Forward primer: 5’-AGGAAACATCCGTCA TGGACT-3’ and Reverse primer: 5’-TAATACGACTCACTATAGGG(T7)ATGGCTTAGGACAGTGTGTGC-3’. WISH was performed according to previously described methods^4–6^. Total RNA was extracted from the embryos at different developmental stages using the Trizol reagent (Invitrogen, Carlsbad, CA, USA), and quantitative real-time PCR was conducted as previously described^7^. The data were analyzed using the △△Ct method, with β-actin used as the house-keeping gene. All experiments were performed in triplicate, and the primer sequences are listed in **Supplementary Table S4**.

#### Construction of a stable cell line with *NARFL* gene knockout was performed in HPMEC cells

Two protein-encoding variants of the NARFL gene were identified from NCBI and Ensemble databases, consisting of 476 and 374 amino acid sequences. The shared exon region of both variants was selected for the design of screening sgRNAs. The location and sequence of the sgRNAs are provided in **Figure 4A in the online-only Data Supplement.**

Wild-type HPMEC cells were collected, and genomic DNA was extracted using the TIANamp Genomic DNA Kit. The *NARFL* target gene was amplified using 2×EasyTaq PCR SuperMix. Five confirmed sgRNAs were used for the construction of sgRNA-Cas9 plasmids. HEK293 cells were cultured and used for virus packaging. After 96 hours, the lentivirus was collected, filtered, and used for cell infection. Genomic PCR amplification was performed to confirm the cleavage effect of the corresponding sgRNAs. The verified cells were then subjected to monoclonal cell selection by seeding one cell per well in a 96-well plate. After two weeks, monoclonal cell communities were selected and expanded into larger plates.

Genomic DNA was extracted from the expanded monoclonal cell lines, and target amplification was performed followed by sequencing to confirm successful gene editing and obtain *NARFL* gene knockout cells. Among the 109 selected monoclonal cells, only four (HPMEC-*NARFL*-sg6-10, 20, 26, 40) showed functional knockout of single alleles. However, HPMEC-*NARFL*-sg6-10 exhibited cell death during the growth process and was deemed unsuitable for further experiments. Therefore, HPMEC-*NARFL*-sg6-20, 26, and 40 were selected for additional verification using Western blot analysis. The results showed that HPMEC-*NARFL* -sg6-40 exhibited the most significant decrease in NARFL expression compared to wild-type cells.

Consequently, HPMEC-*NARFL*-sg6-40 was selected as the final knockout cell line for subsequent experiments, as depicted in **Figure 4D in the online-only Data Supplement.**

#### Ferroptosis Related Indicators Assay

Genomic DNA isolation and genotyping were performed using the NaOH lysis method as previously described^8^. Caudal fins of zebrafish embryos were cut, and the DNA from the tail samples was used for genotyping. The remaining body of the zebrafish embryos was sampled for reactive oxygen species (ROS) assay.

In brief, the embryos were digested with 100 µL of 0.25% (w/v) trypsin/EDTA solution for 10 minutes. The reaction was stopped by adding 200 µL of DMEM containing 10% (v/v) fetal bovine serum (FBS). The sample was then centrifuged at 2500 rpm for 5 minutes at 4°C to remove the supernatant. The cell pellet was washed with 200 µL of PBS containing 2% (v/v) FBS, followed by another centrifugation step. The cells were resuspended in 200 µL of PBS containing 2% FBS and incubated at 37°C for 30 minutes with 10 µM DCFH-DA, BODIPY 493/503 probe, and DPPP probe (Maokang, Shanghai, China). A sample without probe incubation was used as a negative control. The fluorescence was detected using a FACS Canto Flow Cytometer (BD Bioscience, USA) at the excitation/emission wavelengths of 488/525 nm and 351/380 nm.

For the assay using 5 dpf zebrafish embryos cultured at 1×PTU, the embryos were collected and placed in a 24-well cell culture plate with new egg water. A 1 µL solution of BES-H2O2-AC fluorescent dye (1 mg/L, soluble in DMSO), BODIPY 493/503, and DPPP was added to each well with a dilution of 1:10000. The dye solution was gently shaken to disperse it. The plate was then incubated in a 28°C incubator for 2 hours and subsequently observed under a fluorescence microscope.

To inhibit ferroptosis, a 16 μM concentration of Ferrostain-1 (MedChemExpress, LLC, USA) was used for ferroptosis inhibition exposure starting from 24 hpf. After two days of treatment, 3 dpf embryos were collected for ferroptosis measurement.

#### Cell Counting Kit-8 (CCK-8) Cell Proliferation Experiments

Cell proliferation experiments were conducted using the Cell Counting Kit-8 (CCK-8) assay, which utilizes a water-soluble tetrazolium salt, WST-8 (2-(2-methoxy -4-nitrophenyl)-3-(4-nitro phenyl)-5-(2,4-disulfophenyl)-2H-tetrazolium monosodium salt) developed by Dojindo. The WST-8 is reduced by intracellular dehydrogenases in the presence of the electron carrier 1-Methoxy PMS, resulting in the formation of an orange-yellow formazan dye. The amount of formazan dye produced is directly proportional to the number of viable cells and can be measured spectrophoto-metrically.

#### Biochemical Analyses

Cytosolic aconitase activity was assessed using the Aconitase Activity Assay Kit (Sigma-Aldrich, St. Louis, MO, USA). The levels of cellular redox substances, including glutathione (GSH), glutamine, and malondialdehyde (MDA), were measured using colorimetric assays with commercially available assay kits (Beyotime, Nanjing, China). The iron level in zebrafish was determined using a colorimetric assay kit (Dojindo laboratories, Kumamoto, Japan). Vascular endothelial function, including nitric oxide (NO) and endothelin-1 (ET-1), was measured using ELISA assays with commercially available assay kits (LMAI, Shanghai, China).

#### Seahorse Assay

In 5-day-old zebrafish and HPMEC cells (20,000 cells/well), the oxygen consumption rate and extracellular acidification rate (a surrogate marker of glycolysis) were measured using an XFe24 Extracellular Flux Analyzer (Seahorse Biosciences). Sequential addition of 1μM Oligomycin, 0.5 μM FCCP, and 2 μM Rotenone plus 0.5 μM Antimycin was performed, as previously described^9^.

#### *In vitro* Angiogenesis Assays

Tube formation was evaluated using a commercial kit, In vitro Angiogenesis Assay Kit (Chemicon International, Temecula, USA). Matrigel with reduced growth factors (100 μl/well) was pipetted into a pre-chilled 48-well plate and polymerized at 37℃ for 30 min. HPMECs and HPMEC-*NARFL-/-* cells treated differently (2×10^5^ cells/well) were suspended in 100 μl of basic media and seeded onto the Matrigel-coated plate. After incubation for 4-6 hours, tubular structures were photographed using an Olympus microscope at 20× magnification. The acquired images were then analyzed using the angiogenesis analysis plugin in Image J software for node count, intersection count, mesh number, mesh area, vascular branch count, total vascular length, vascular branch length, and trunk length. The number of branch points was determined by quantifying triplicate determinations from three separate experiments.

#### The Ciao3+/- mouse model was generated using the CRISPR/Cas9 method

A Ciao3 (NARFL homologous) hybrid was constructed in C57BL/6 mice, and the mouse model was successfully knocked out. The Ciao3 gene motif (NCBI: NM_026233.8; Chromosome 17 Ensembl: ENSMUSG000 00002280) in mice consists of a total of 11 exons (transcript Ciao3-201: ENSMUST0 0000002350), with exons 3-4 selected as the knockout target. Exon 3 starts from approximately 11.41% of the coding region, and exons 3-4 account for 19.4% of the coding region, resulting in an effective knockout region size of 1820 bp. A combination of Cas9 and guide RNA (gRNA) was injected into fertilized eggs, resulting in targeted knockout of the offspring. The resulting F0 generation was screened using PCR, and wild-type mice were bred to confirm germline transmission and produce F1 offspring. Heterozygous mice were then mated to generate homozygous generations. Gene identification was performed using the following primer sequences: Primer 1: F1: 5’-CTGGCTCAGACCATTTCTGCATC-3’; R1: 5’-GTGATGCTGCCA AACACTCGTCA-3’. The wild-type fragment size was 2509 bp, and the mutant fragment size was 683 bp. Primer 2: F1: 5’-CTGGCTCAGACCATTTCTGCATC-3’; R1: 5’-TTTTCTATTTCCTGA CAGTA GGTGG-3’. The wild-type fragment size was 523 bp, and the heterozygous fragments were identified as follows: a fragment size of 683 bp with primer 1, and a fragment size of 523 bp with primer 2.

#### Immunoblotting and Co-IP

For immunoblotting, cells were lysed in Laemmli buffer, and the protein lysates were resolved by SDS-PAGE and transferred onto a PVDF membrane. The membranes were then blocked in 5% non-fat milk or BSA in PBS buffer with 0.1% Tween (PBST) and incubated overnight at 4 ℃ with primary antibodies. After washing with PBST buffer, the membranes were incubated with secondary antibodies for 1 hour at room temperature. Immunoreactive bands were visualized using the enhanced chemiluminescence (ECL) system.

The primary antibodies used were: NARFL (NOVUS, 1:1000), GPX4 (NOVUS, 1:1000), TFR (PK17158, 1:500), Ferritin (T55648, 1:1000), IRP1 (T55075, 1:1000) from Abmart, SLC7A11 (A2413, 1:2000) from CST, CYP2J2 (ATA27790, 1:2000) from Atagenix, NARFL (sc-514078, 1:500), MMS19 (sc-390028, 1:500), FAM96B (sc-376801, 1:5000), and CIAO1 (sc-374498, 1:500) from Santa, and GAPDH (ab8245, 1/6000) from Abcam.

For the mouse embryo, fine fragments were obtained and lysed at a ratio of 200 µL per 20 mg of tissue. The lysate was homogenized and then centrifuged at 4 ℃ at 12000g for 15 minutes. The supernatant was collected, and the rest of the procedure was the same as for the cells.

For Co-IP, Protein A/G microspheres were washed with PBS and prepared as a 50% Protein A/G working solution. About 1 μg of IgG of the same species as the IP monoclonal antibody and 100 μL of Protein A/G working solution were added to 1 mL of cleavage solution. The mixture was incubated at room temperature for 1 hour and then centrifuged at 13,000 g for 10 minutes. The supernatant was transferred to a new centrifuge tube to remove non-specific binding of proteins to immunoglobulins. Then, a specific volume of antiprecipitation antigen was added, and 100 μL of Protein A/G working solution was added to capture the antigen-antibody complex. The mixture was incubated overnight at 4 ℃ with agitation. After centrifugation, the precipitation was collected and washed with pre-cooled PBS three times. The supernatant was removed by centrifugation, and the precipitation was retained. The precipitation was re-suspended in 100 μL of 1× loading buffer and boiled in water at 100 ℃ for 5 minutes. Before loading, all samples were centrifuged at 4 ℃ at 13,000 g for 10 minutes, and the samples were loaded at 20 μL per well.

#### Immunohistochemistry and Immunofluorescence of Lung and Mouse Embryo Sections

Cryostat sections were prepared from 5 µm thick OCT-embedded lung tissues and mounted on gelatin-coated histological slides. The slides were left to thaw at room temperature for 20 minutes and then rehydrated in wash buffer for 10 minutes. All sections were blocked using 10% goat serum and incubated with primary antibodies overnight at 4°C, followed by incubation with Alexa 488, CY3, and CY5-conjugated secondary antibodies (Thermo Fisher Scientific) for immunofluorescence. Primary antibodies against NARFL (NBP1-83611, 1:200) and CD31 (ab182981, 1:100) were obtained from Novus Biologicals and Abcam, respectively. Primary antibodies against α-SMA (BM0002, 1:100) and CYP2J2 (ATA27790, 1:500) were purchased from Boster Biological Technology and Atagenix, respectively. Primary antibodies against Endomucin (GB112648, 1:300), CD34 (GB13584, 1:200), γH2AX (GB111841, 1:200), CD31 (GB113151, 1:200), and SMA (GB13044, 1:1000) were obtained from Servicebio. Imaging was performed using a Leica confocal microscope (TCS SP8). Small pulmonary vessels (<100 μm diameter) without association with bronchial airways were selected for analysis in each tissue section (>10 vessels/section). The intensity of staining was quantified using ImageJ software (NIH). The degree of pulmonary arteriolar muscularization was evaluated in OCT lung sections stained for α-SMA by calculating the proportion of fully and partially muscularized peripheral (<100 μm diameter) pulmonary arterioles, as described previously^10^. CD31, an endothelial cell marker, and CD34, an endothelial progenitor cell marker, were used for immunofluorescence detection of whole embryos, and DNA damage in the embryos was assessed by γ-H2AX detection.

#### *Ex vivo* Mice Aortic Ring Assay

The subpackaged matrix glue was thawed at 4 ℃, and 100 μL of the glue was spread in each well of a 48-well cell culture plate. The plate was then incubated in a 37 ℃ incubator for 30 minutes to allow the matrix glue to solidify. Three multiple holes were created in each well. Under sterile conditions and after ether anesthesia, aortas were extracted from both wild-type and Ciao3+/- mice. Para-aortic fat and other tissues were carefully removed, and the aortas were divided into rings approximately 1mm wide. These aortic rings were placed on top of the solidified matrix glue. An additional 100 μL of melted matrix glue was added to cover the aortic rings, and the culture plate was incubated in a 37 ℃ incubator for 30 minutes. Then, 200 μL of ECM medium containing 5% FBS was added, and the cultures were maintained for 4 days. After the incubation period, images were captured under a microscope, and the acquired images were analyzed using the angiogenesis analysis plug-in in ImageJ. The analysis included quantification of the number of blood vessel branches, total length of blood vessels, and length of blood vessel branches.

#### The NARFL gene’s tagSNPs were detected using the SNaPshot method

Initially, the NCBI website (<u>NCBI.nlm.nih.gov</u>) was accessed and a search for the “NARFL” gene was conducted, focusing on the Homo sapiens results. The position of the *NARFL* gene, 79765-79099, was identified. The VCFtoPed tool was then utilized to acquire variation data for this gene in the Chinese population from the Homo sapiens section of the Ensembl database (http://grch37.ensembl.org/Homo_sapiens/). Mutagenesis was conducted using the Haploview 4.2 software, with the linkage format being selected. Following data importation, the marker check interface appeared, where parameters such as the Haploview balance cutoff value and MAF (Minor allele frequency) cutoff value were set. Upon filtering and selecting functional SNPs, the Tagger function was employed to screen the markers based on the selected SNPs, using an r^2^ threshold of 0.8. The outcome of this screening resulted in the selection of 49 TagSNPs. For reference, the Ensembl database was accessed again (http://grch37.ensembl.org/Homo_ <u>sapiens./Gene/VariationGene/Tabledb=core;g=ENSG00000103245</u>;r=16:779753-791329), and 49 TagSNPs were further shortlisted based on mutation type and 10 case-control preliminary experiments. Finally, seven TagSNPs (rs61112891, rs2071952, rs117952680, rs9928077, rs3752556, rs11248948, and chr-731143) were chosen for further analysis using large sample sizes. The SNP detection utilized the SNaPshot method, which follows the dideoxy termination principle of direct DNA sequencing. However, only fluorescently labeled ddNTPs corresponding to specific SNPs were used. By designing sequencing primers in close proximity to the SNP site, multiple SNP sites can be detected simultaneously. This involves DNA extraction, sample sorting, DNA detection, primer synthesis, PCR amplification, alkaline phosphatase treatment in a PCR station, and sequencing using an ABI 3730 XL sequencer.

## Supplemental Figures and Figure Legends

**Supplemental Figure 1.**
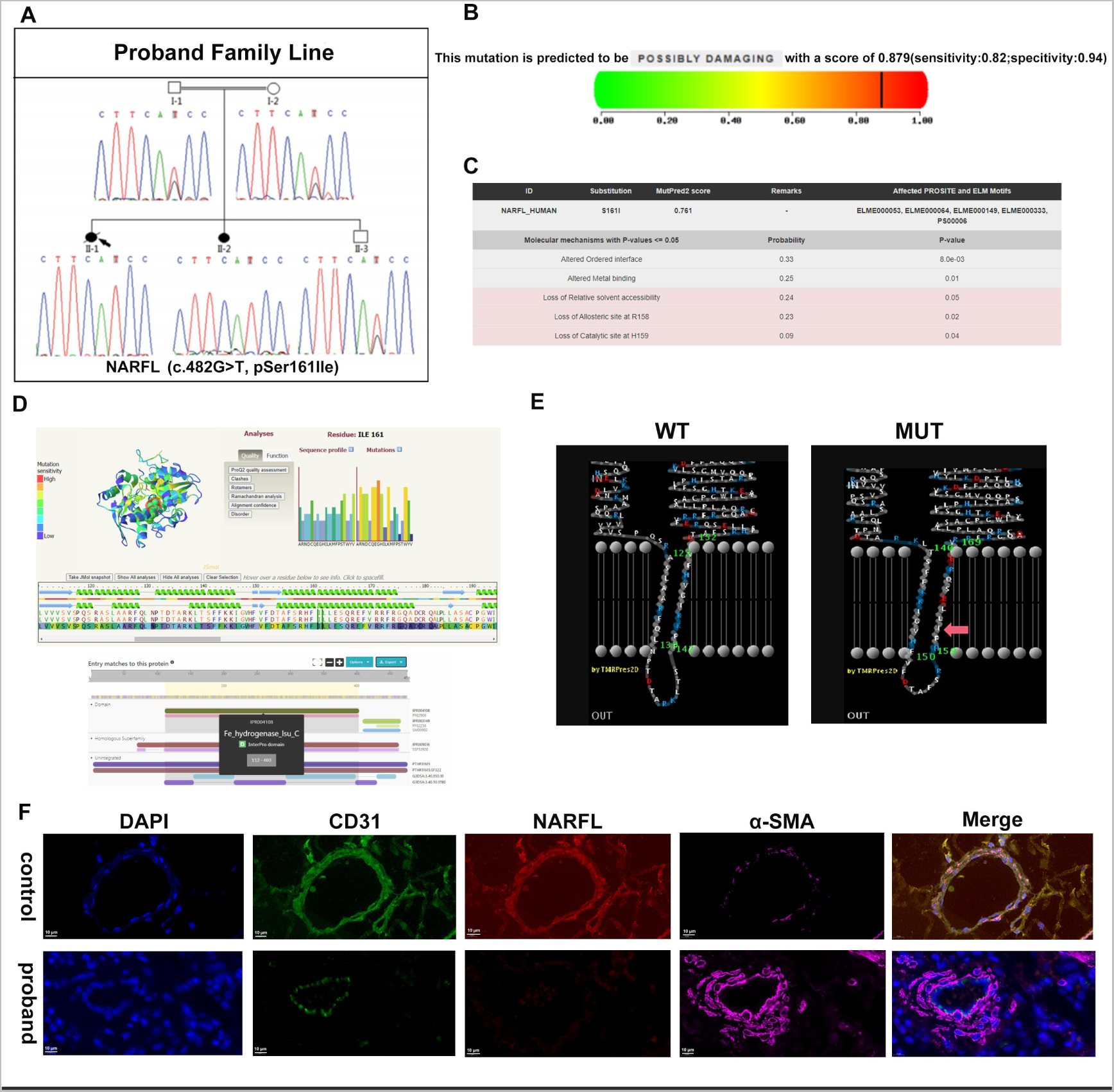
Family with Pulmonary Hypertension Secondary to Diffuse Pulmonary Arteriovenous Malformation. (A) Sequencing results from the family members with pulmonary hypertension secondary to diffuse pulmonary arteriovenous malformation show a homozygous mutation in exon 5 of the NARFL gene. The mutation involves a change from AGC to ATC (hg19NM_002493 c.482 G>T) resulting in an amino acid substitution from serine (Ser, S) to isoleucine (Ile, I). (B) The pathogenicity of the mutation was predicted using Polyphen 2 software, which indicates that it is a deleterious mutation. (C) The VarSome software predicts the effects of the mutation site, suggesting that it may alter the metal-binding domain. (D) The Phyre2 software was used to predict the functional implications of the mutation region (c.482 G>T), indicating that the T mutation is located in the functional region of the ferric hydrogenase. (E) The PRED-TMBB software predicted a change in the mutation region from a non-transmembrane region to an intra-transmembrane region. (F) Immunohistochemical staining of lung tissue from the affected family members with pulmonary hypertension secondary to diffuse pulmonary arteriovenous malformation. The staining shows the presence of CD31 (green) expressed in endothelial cells, α-SMA (pink) expressed in myofibroblasts, and NARFL (red) expressed in the tissue cytoplasm. Merge images show the combination of all staining results with a field of view magnification of 100×.

**Supplemental Figure 2.**
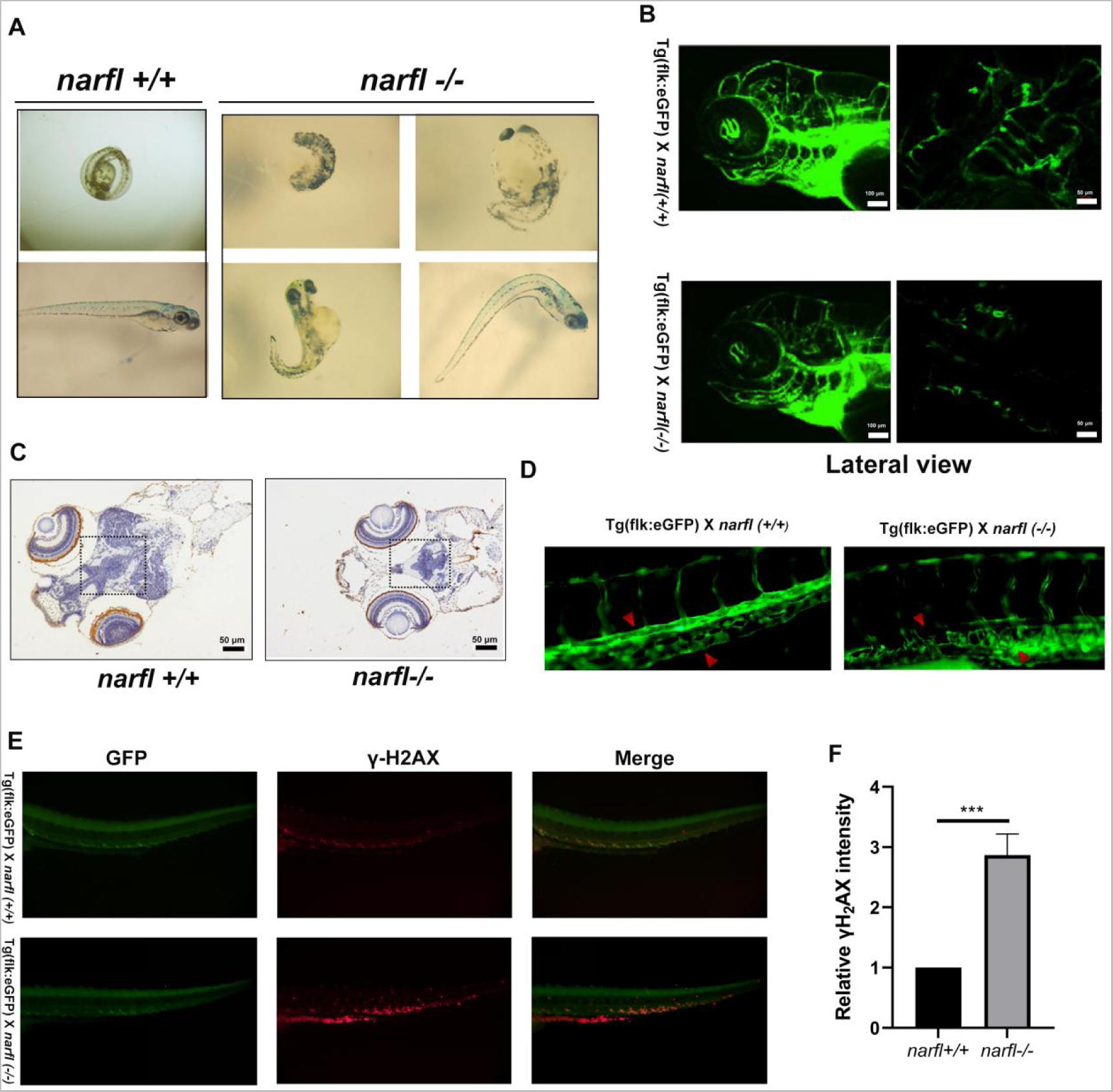
Deletion of the *narfl* gene Causes Abnormal Development and Vascular Structure in Zebrafish. (A) Morphological deformities during development were observed in *narfl-/-* zebrafish. (B) Fluorescence inverted microscope imaging system was used to observe the blood vessels in the lateral field of the zebrafish brain with different genotypes. (C) TUNEL staining of paraffin sections of the zebrafish brain at 9 dpf with different genotypes showed DNA damage. The field of view was observed at 100× magnification. (D) Fluorescence confocal microscope observation and quantitative analysis of zebrafish vascular segments showed obvious disorganization in the structure of the dorsal aorta and posterior main vein in *narfl-/-* zebrafish. (E-F) The γH2AX test revealed significant DNA damage in the dorsal aorta and portions of the posterior main vein. Statistical analysis of the results showed significant differences compared to the control groups: \**p*<0.05, \*\*\**p*<0.001.

**Supplemental Figure 3.**
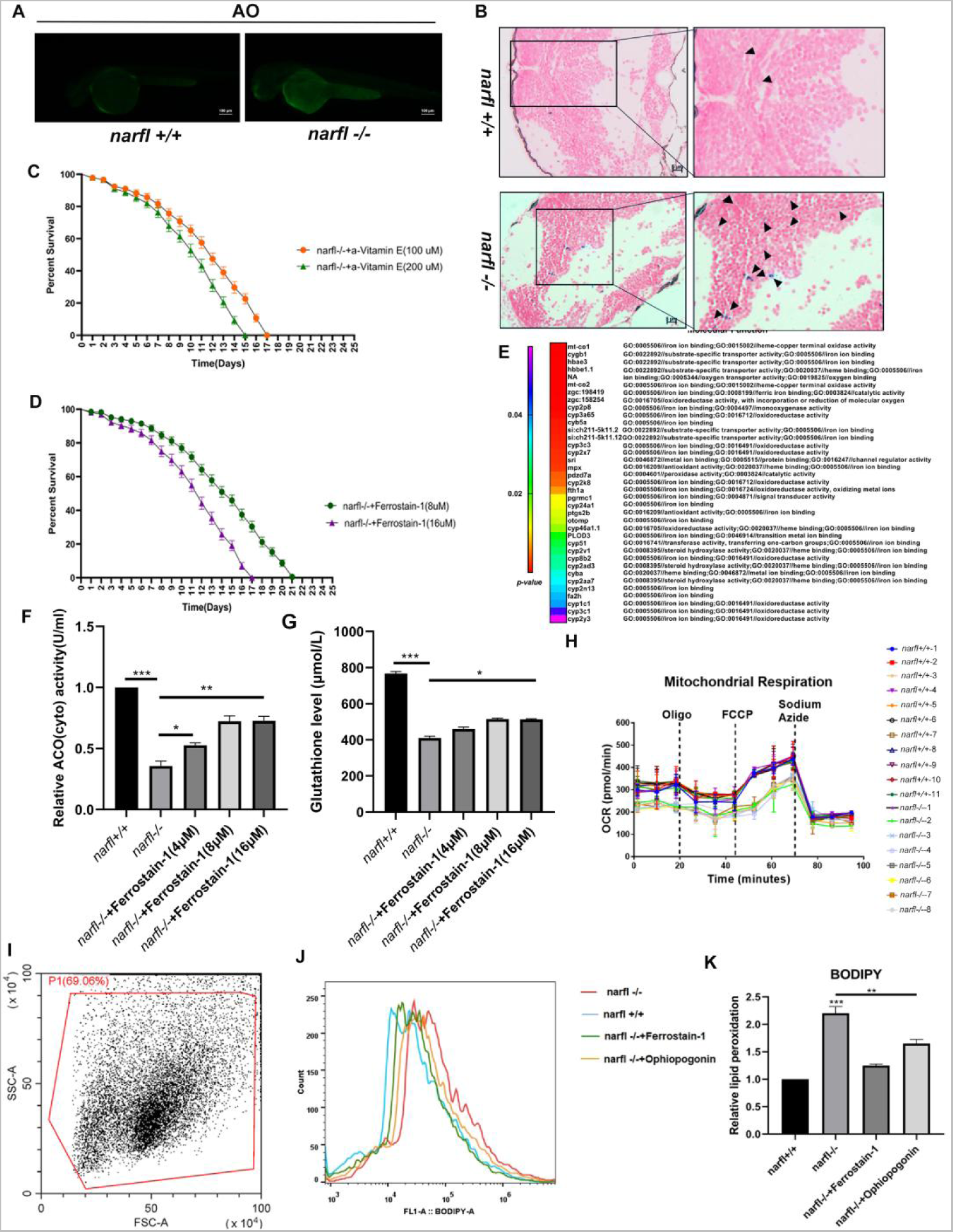
n*a*rfl Gene Deletion Induces Oxidative Stress, Lipid Peroxidation, and Iron Level Increase in Zebrafish. (A) Apoptosis level in 5-day-old zebrafish was assessed by AO staining. Green fluorescence intensity reflects the apoptosis level, and the visual field magnification was set as 200×. (B) Hemosiderin content in the brain of 7-day-old zebrafish was determined by Prussian blue staining. The blue region indicated by the black arrow represents the positive part, with visual field magnifications set at 200× and 400× respectively. (C) Zebrafish embryos were treated with α-Vitamin E at concentrations of 100 μM and 200 μM. The survival time of *narfl-/-* zebrafish was extended to 15 dpf with 200 μM α-Vitamin E, and to 17 dpf with 100 μM α-Vitamin E. (D) Zebrafish embryos were treated with Ferrostain-1 at concentrations of 8 μM and 16 μM. The survival time of *narfl-/-* zebrafish was extended to 17 dpf after 16 μM Ferrostain-1 treatment, and to 21 dpf after 8 μM Ferrostain-1 treatment. (E) List of iron metabolism-related genes differentially expressed in transcriptome sequencing between wild and *narfl-/-* zebrafish. (F) Cytoplasmic cis-aconitase activity was measured in zebrafish at 5 dpf after treatment with Ferrostain-1 at concentrations of 4 μM, 8 μM, and 16 μM. (G) Glutathione levels in zebrafish were assessed at 5 dpf after treatment with Ferrostain-1 at concentrations of 4 μM, 8 μM, and 16 μM. (H) Mitochondrial respiration was evaluated in 11 wild-type and 8 *narfl-/-* zebrafish. (I-K) BODIPY levels in wild-type and *narfl-/-* zebrafish treated with Ferrostain-1 and Ophiopogonin were measured by flow cytometry. \**p*<0.05, \*\**p*<0.01, \*\*\**p*<0.001.

**Supplemental Figure 4.**
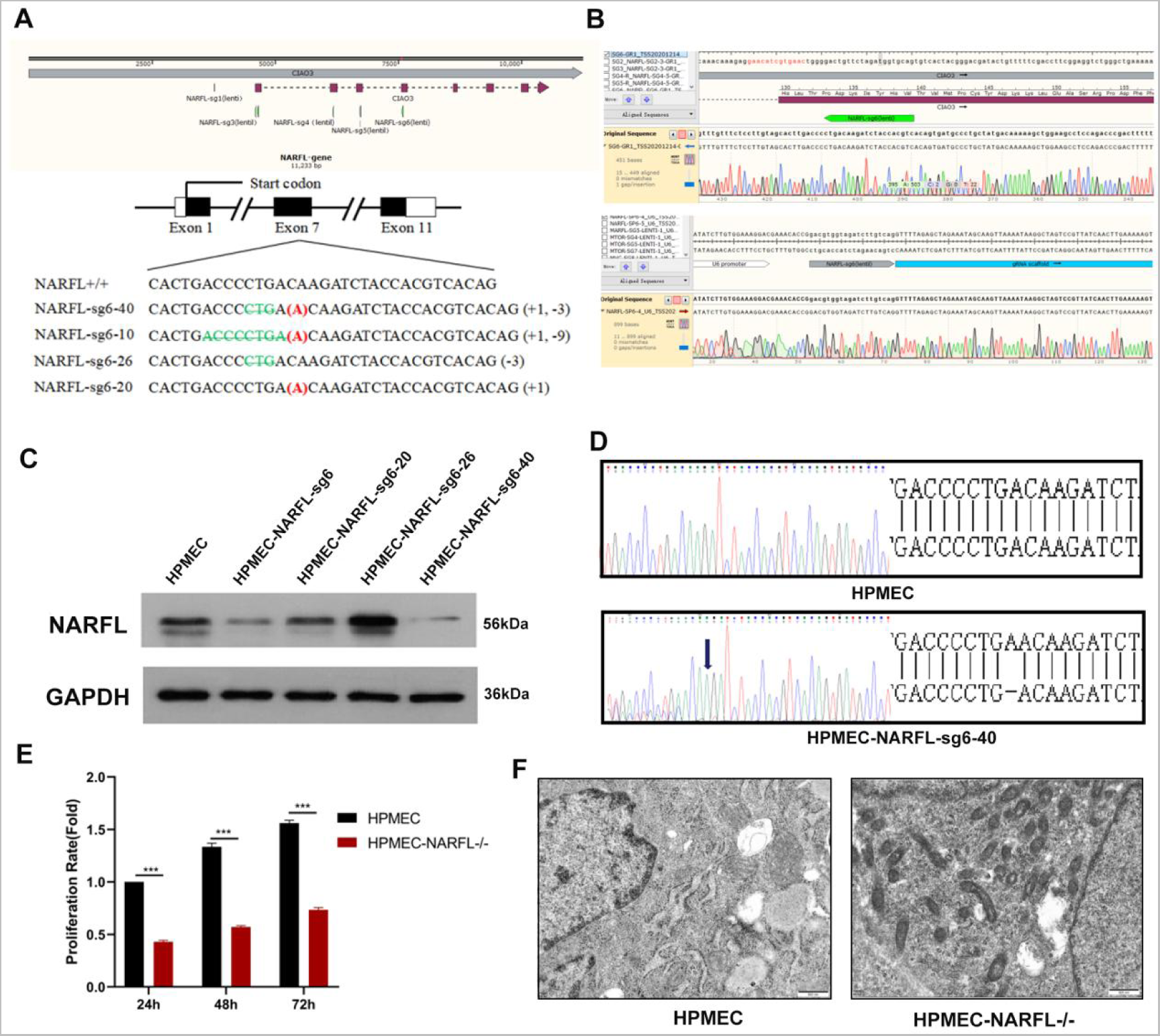
Construction of NARFL Gene Knockout Stable Cell Line in HPMEC Cells. (A) Design location of sgRNA. (B) Plasmid sequencing results. (C) Western blot analysis of monoclonal protein. Compared to wild-type cells, NARFL expression was significantly down-regulated in HPMEC-NARFL-sg6. NARFL expression in HPMEC-NARFL-sg6-20 was comparable to wild-type cells. NARFL expression was higher in HPMEC-NARFL-sg6-26 compared to wild-type cells. NARFL expression was significantly down-regulated in HPMEC-NARFL-sg6-40. (D) DNA sequences of HPMEC and HPMEC-NARFL-sg6-40. (E) Cell proliferation capacity was quantitatively determined using the CCK-8 method at 24h-72h. \*\*\**p*<0.001. (F) Abnormal morphology of endothelial mitochondria due to NARFL gene deletion observed by electron microscope.

**Supplemental Figure 5.**
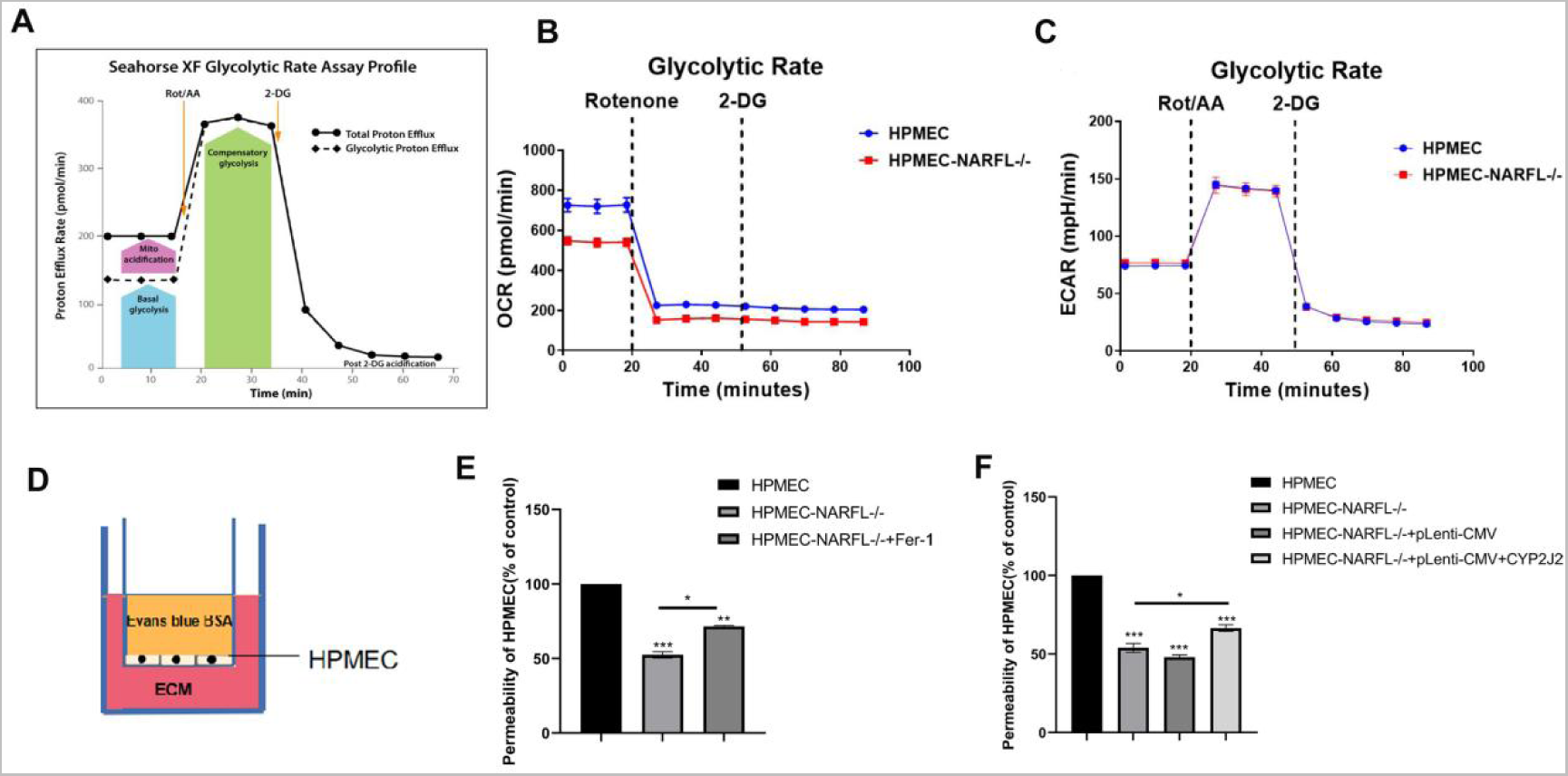
Effect of NARFL Gene Deletion on Endothelial Cell Glycolysis and Endothelial Dysfunction. (A) Model diagram illustrating the calculation of key parameters of glycolysis in the cell glycolysis rate curve. The basic proton flow rate represents the number of protons released by cells into the detection solution before the addition of rotenone or antimycin A. The mitochondrial acidification rate is the product of the mitochondrial oxygen consumption rate and carbon dioxide contribution coefficient. Basal glycolysis is the difference between the basal proton flow rate and mitochondrial acidification rate. Compensatory glycolysis refers to the highest proton outflow rate after the addition of rotenone or antimycin A. Acidification after the addition of 2-DG refers to the lowest value of proton flow rate after the addition of 2-DG. (B) Glycolysis rate curves of HPMEC and NARFL mutated HPMEC cells. Blue represents wild-type HPMEC, and red represents NARFL mutated HPMEC. (C) Results of extracellular acidification showed no significant difference. (D) Diagram of the Evans Blue cell penetration experiment. Cells were inoculated in the Transwell chamber, and ECM medium was placed in the lower chamber. (E-F) Quantitative results of the Evans Blue cell penetration experiment.

**Supplemental Figure 6.**
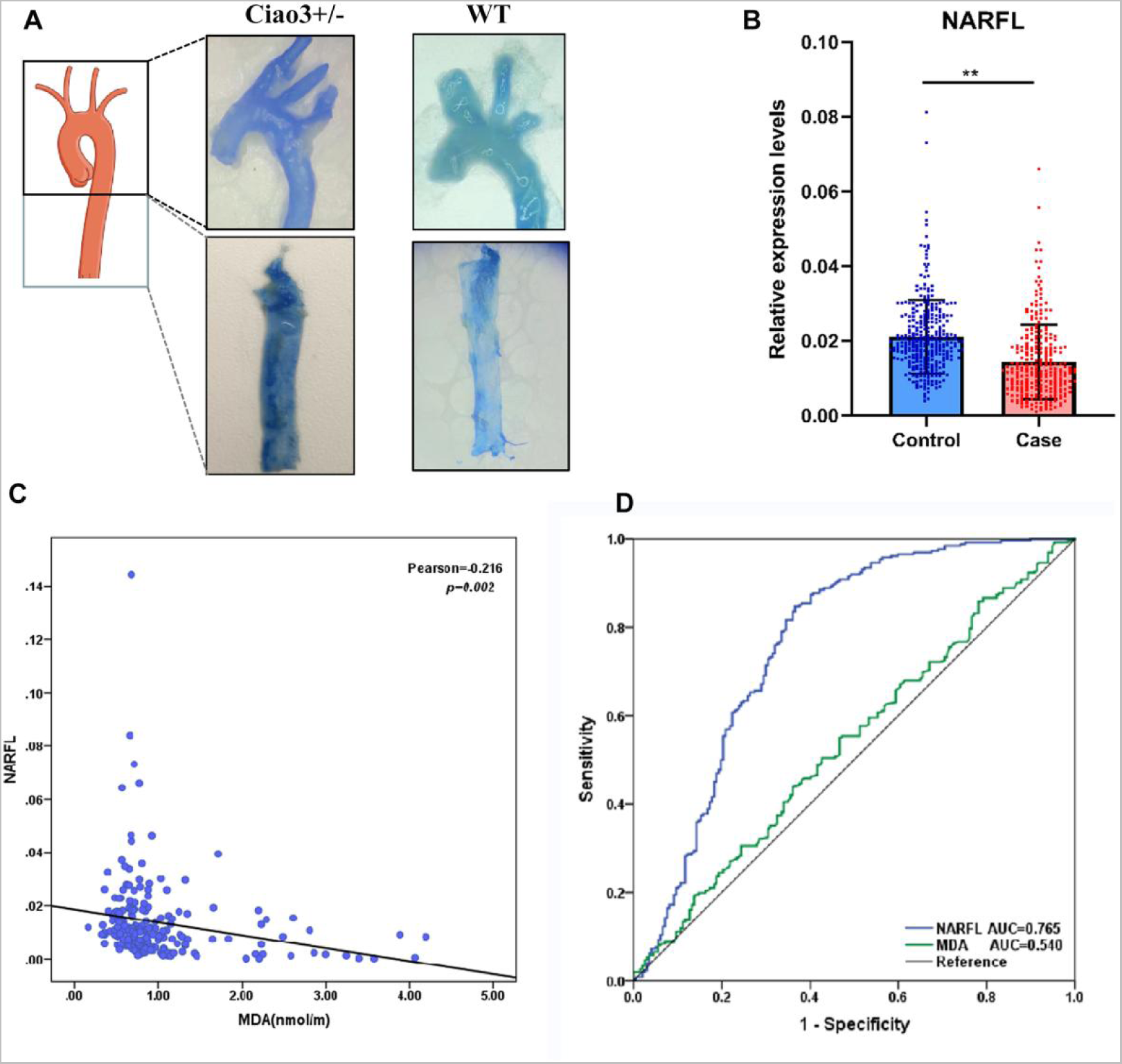
Impairment of Vascular Function in Ciao3 Heterozygous Mice and NARFL Polymorphisms as Susceptible Sites for Vascular Endothelial Dysfunction Diseases. (A) Evans Blue staining results of the aortic arch in Ciao3+/- mice showed significantly darker staining compared to the wild-type group, indicating vascular dysfunction. (B) NARFL expression levels were significantly lower in the disease group, \*\**p*<0.01. (C) The expression of NARFL was negatively correlated with MDA levels. (D) Receiver Operator Characteristic (ROC) curve was generated to compare the diagnostic ability of the MDA levels and NARFL expression levels in the disease population with tagSNP difference. The results demonstrated that NARFL expression levels provided better discrimination of the population with tagSNP difference. The area under the curve (AUC) for NARFL was 0.765, while the AUC for MDA was 0.540, indicating a lower discriminative ability compared to NARFL.

